# Risk-benefit profile of edoxaban and warfarin in patients with atrial fibrillation: a comprehensive systematic review and meta-analysis of randomized trials

**DOI:** 10.1101/2025.03.05.25323458

**Authors:** Muhammad Ahmad Sohail, Asna Moghis, Muhammad Waleed Imran, Misha Khalid, Sanwal Sardar Nawaz, Muhammad Hamza Akram, Asaad Akbar Khan

## Abstract

**Introduction:** Atrial fibrillation is the most prevalent cardiac arrhythmia in the United States and substantially increases the risk of stroke and heart failure in patients with non-valvular atrial fibrillation, those undergoing percutaneous coronary intervention, and transcatheter aortic valve repair. Warfarin, a vitamin K antagonist, has long been the standard anticoagulant therapy but is limited by drug interactions and monitoring requirements, while edoxaban, a factor Xa inhibitor, has shown favorable bleeding outcomes.

**Methods:** This meta-analysis included five randomized controlled trials identified through a comprehensive search of PubMed, Medline, Embase, Google Scholar, CENTRAL, and ClinicalTrials.gov from January 2014 to October 2024 (PROSPERO ID: CRD420250648890). Data were pooled using the inverse variance method with hazard ratios and 95% confidence intervals, DerSimonian–Laird random-effects model was applied in Stata version 17. Risk of bias was assessed using the Cochrane RoB v2 tool and certainty of evidence using GRADE assessment tool, with statistical significance set at P < 0.05.

**Results:** Edoxaban demonstrated a greater reduction in stroke or systemic embolism overall but was less effective in patients with prior myocardial infarction (HR 0.58, 95% CI 0.32–1.05). Major adverse cardiovascular events marginally favored edoxaban over warfarin (HR 0.90, 95% CI 0.83–0.98, P = 0.01), whereas warfarin performed better in patients with CHA₂DS₂-VASc scores ≥4. No significant difference was observed in all-cause mortality (P = 0.32). For clinically relevant non-major bleeding, warfarin was more favorable in patients aged ≥65 years and those with normal renal function, while edoxaban reduced major bleeding more effectively in patients with <25% heart failure; warfarin showed slight superiority when heart failure prevalence was ≥25%.

**Conclusion:** Overall, edoxaban shows robust efficacy across multiple outcomes, while warfarin remains essential in specific high-risk subgroups, warranting further confirmatory studies.

## Introduction

The most common arrhythmia worldwide is Atrial Fibrillation (AF), a major risk factor for common comorbidities such as stroke, dementia, and heart failure.[1] Unfortunately, its numbers are on the rise, with a predicted increase in cases reaching 12 million in the United States alone by 2030. This trend can be credited to changing lifestyle patterns and the increasing elderly population. [2,3]

To treat AF efficiently, it is important to understand its two primary causes. The first, Valvular AF, is mainly associated with mitral stenosis or artificial heart valves. The second, non-valvular type can be attributed to all other forms of valvular heart disease. [4,5] Procedures such as transcatheter aortic valve replacement (TAVR) and percutaneous coronary intervention (PCI) may also cause new onset AF (NOAF), by causing increased ectopic activity within atrial muscles. [6,7,8] This cause is particularly concerning as 13% of TAVR patients develop NOAF and have a higher risk of stroke, Myocardial infarction, and mortality as compared to those with pre-existing AF. [9,10]

The most pivotal therapy for AF to date has been anticoagulants. A widely known anticoagulant is Warfarin, which works by inhibiting hepatic synthesis of clotting factors (II, VII, IX, and X).[11] By significantly reducing clot formation and hence the risk of stroke in NVAF patients, Warfarin was considered the gold standard for a long time.[12] However, along with its benefits come several drawbacks, including its narrow therapeutic range and frequent drug interactions.

Moreover, strict INR monitoring became necessary to prevent toxicity and bleeding.[13]

In order to conquer these obstacles, Asinger et al. emphasized substitute therapeutic agents such as direct oral anticoagulants (DOACs) having similar efficacy to warfarin. However, variable safety of different DOACs depending on the type of drug and its dose were reported particularly in the.elderly population. This requires critical consideration in choosing the most appropriate anticoagulation therapy. [14]One factor Xa inhibitor, edoxaban, a promising DOAC, has demonstrated its efficacy and safety equivalent to warfarin. The key trial ENGAGE AF-TIMI 48 trial demonstrated that both drugs had comparable effects in stroke prevention and systemic embolism (SSE), with edoxaban indicating much reduced percentages of hemorrhage and heart-related death. [15,16, 17]

Our detailed meta-analysis highlights the importance of opting for the suitable anticoagulation among patients with NVAF, including. those that received PCI and TAVR, based on various key indicators to improve patient’s survival and mortality. Our synthesis of evidence based on a variety of studies provides an in-depth analysis and understanding of the efficacy and safety profiles of edoxaban and warfarin.

## Methods

### Data Sources and Searches

The meta-analysis comprised five studies that were located subsequent to a systematic review of PubMed, the Embase, Google scholar, ClinicalTrials.gov, and Cochrane Central Register of Controlled Trials (CENTRAL). The search strategy has been entered on PROSPERO (CRD420250648890).The search keywords and Medical Subject Headings (MeSH) were edoxaban, warfarin, atrial fibrillation, efficacy and safety. The search was optimized with the help of Boolean operators (AND, OR). There were no restrictions of language. Also, the screening of the relevant articles and systematic reviews was performed manually in order to further identify the eligible studies.

#### Selection Criteria

One criterion was used that incorporated the following 1) We included studies that report specific data on 5 outcomes: Major Adverse Cardiac Events, stroke or systemic embolism (S/SE), all-cause mortality, major bleeding and clinically relevant non-major bleeding. 2) Edoxaban and warfarin clinical trials in the atrial fibrillation patients. 3) Reporting at least one outcome of interest, e.g. stroke and systemic embolism, major bleeding, all-cause mortality, clinically relevant non-major bleeding, major adverse cardiac events or serious adverse effects. Further, we excluded studies that: 1) were Non-comparative, a case report, observational, and/or editorials. 2) Patients who were enrolled and had other indications than atrial fibrillation. 3) Lead to inadequate or redundant data. Two reviewers examined the title and abstract of the identified studies and then evaluated eligibility by full-text. When discrepancies were identified, a discussion or consultation with a third reviewer was done to resolve them.

### Data Mining and Acuity Evaluation

The identified data were collected in a standardized data extraction sheet where the necessary information was provided such as the characteristics of the study (author, year, design, sample size, follow-up duration), patient characteristics, intervention characteristics (dosage, administration), and clinical outcomes. Two independent reviewers extracted the data. The risk of bias on included RCTs was evaluated based on the revised Cochrane Risk of Bias tool (RoB 2.0) [18], which assesses the following domains randomization, deviations of planned interventions, the absence of outcome data, outcome-measurement, and selective reporting.

Each outcome has been evaluated based on the quality of provided evidence using the Grading of Recommendations, Assessment, Development, and Evaluations (GRADE) framework. [19] The duplicates of the study were eliminated using EndNote X7 software.

### Data Synthesis and Analysis

StataCorp. (2021) was used to conduct a meta-analysis. Stata: Release 17. StataCorp LLC, Texas, USA. DerSimonian-Laird random-effects model [20] and the inverse-variance approach was applied to all the studies. The estimates of the effects were in the form of Hazard Ratio (HR) with 95% confidence interval (CI). The I2 statistics, [21] was used to measure heterogeneity with 25, 50 and 75 percent values representing low, moderate and high heterogeneity respectively. They were divided into subgroups, which were analyzed by key clinical characteristics, such as CHA2DS2-VASc score [score>=4, high risk; score<4=moderate risk], age, renal clearance (CrCl), history of myocardial infarction (MI) and heart failure (HF) where percentage prevalence of the four outcomes were regarded in the subgroups respectively. The test by Egger was also carried out to test the publication bias on a rigorous basis.

## Results

This meta-analysis involved 26,832 participants in five clinical trials in order to compare the effectiveness and safety of edoxaban and warfarin in patients with atrial fibrillation. The chosen articles differed in terms of the characteristics of the populations, duration of treatment, and the definition of outcomes, yet all of them fulfilled the inclusion criteria to make the systematic and comprehensive comparison of the two anticoagulants. Figure 1 shows that a PRISMA flowchart displays the criteria. Major adverse cardiac events (MACE), stroke and systemic embolism (SSE), all-cause mortality (ACM), major bleeding (MB), and clinically relevant non-major bleeding (CRNM) were the key outcomes [under analysis] Table 1 indicates baseline characteristics.

**Table 1.** Baseline characteristics.

| Study name and Year | Type of study | Total no. of patients | Dosage of Edoxaban (mg) | Sex (%) | Mean age (years) | Mean BMI (kg/m<sup>2</sup>) |
| --- | --- | --- | --- | --- | --- | --- |
| **Bergmark 2024** | RCT | 21105 | 60/30 | F = 38.1<br>M = 61.9 | 70.5 | - |
| **Meighem 2021** | RCT | 1426 | 60 | F/M = 48.7 | 82.1 | 27.7 |
| **Goette 2020** | RCT | 2199 | 60 | F = 34.4<br>M = 65.6 | 64.3 | 30.7 |
| **Sankyo 2020** | RCT | 1500 | 60/30 | F = 25.8<br>M = 74.2 | 69.4 | - |
| **Hohnloser 2019** | RCT | 602 | 60/30 | F = 29.1<br>M = 70.9 | 60.5 | 28.1 |

RCT = Randomized controlled trials; F = Female; M = Male

Figure 1. PRISMA flowchart

### Major adverse cardiac events (MACE)

The combined analysis showed that edoxaban had a major decrease in the risk of MACE as compared to warfarin (HR 0.90, 95% CI 0.83-0.98, p = 0.01). The heterogeneity was low (I2 = 0.00%). This implies that edoxaban can be a more suitable choice in the prevention of major cardiovascular events. Nonetheless, there was a difference in the benefit of this based on the subgroup analysis depending on patient profile. The same trend was observed in patients with preserved renal functioning (CrCl 80 mL/min) in favour of edoxaban. On the other hand, warfarin, which has been thought to be a more effective deterrence of MACE in patients with a CHA 2 DS VASc score of 4 or above, emerged as a potentially significant participant in the prevention of cardiovascular events in patients with a higher thromboembolic risk. The effect of heart failure status on MACE outcomes was also assessed in which patients with substantial burden of HF were more likely to respond better on warfarin as compared to patients with little or no HF which would respond better with edoxaban. This was shown in Figures 1-7.

**Figure 1.**
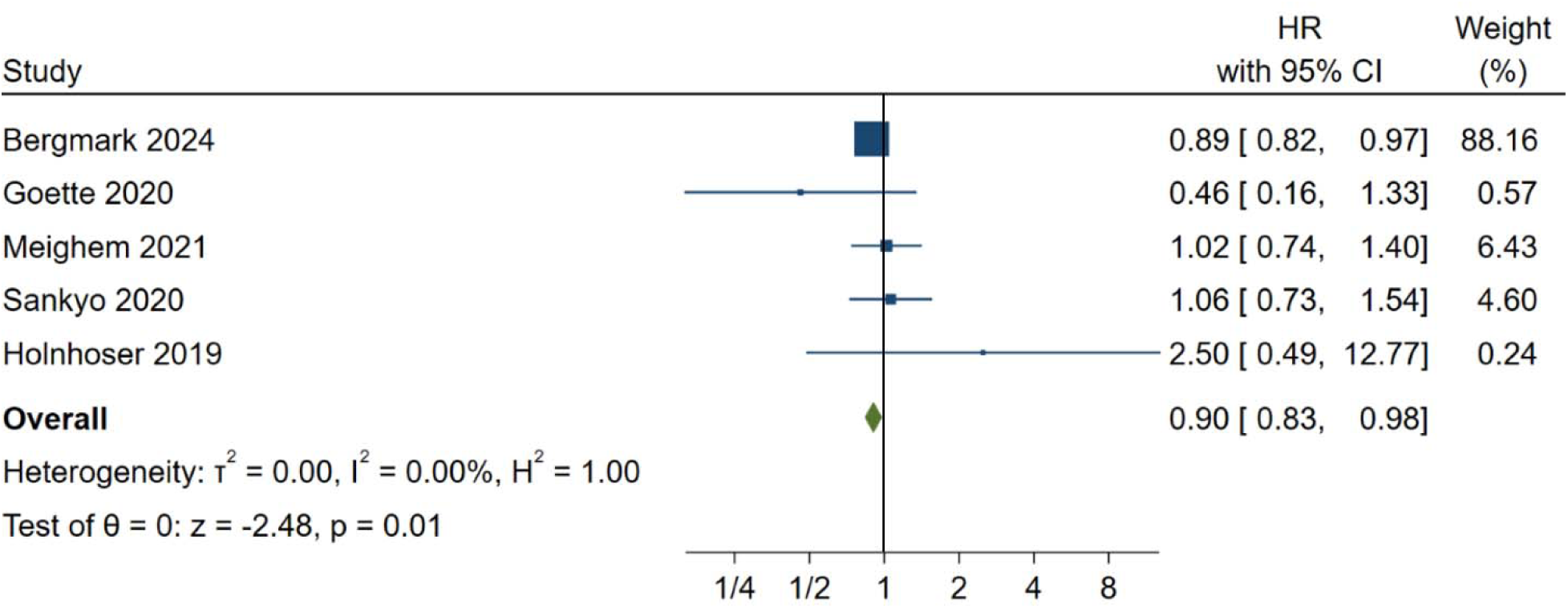
Major adverse cardiac events

**Figure 2.**
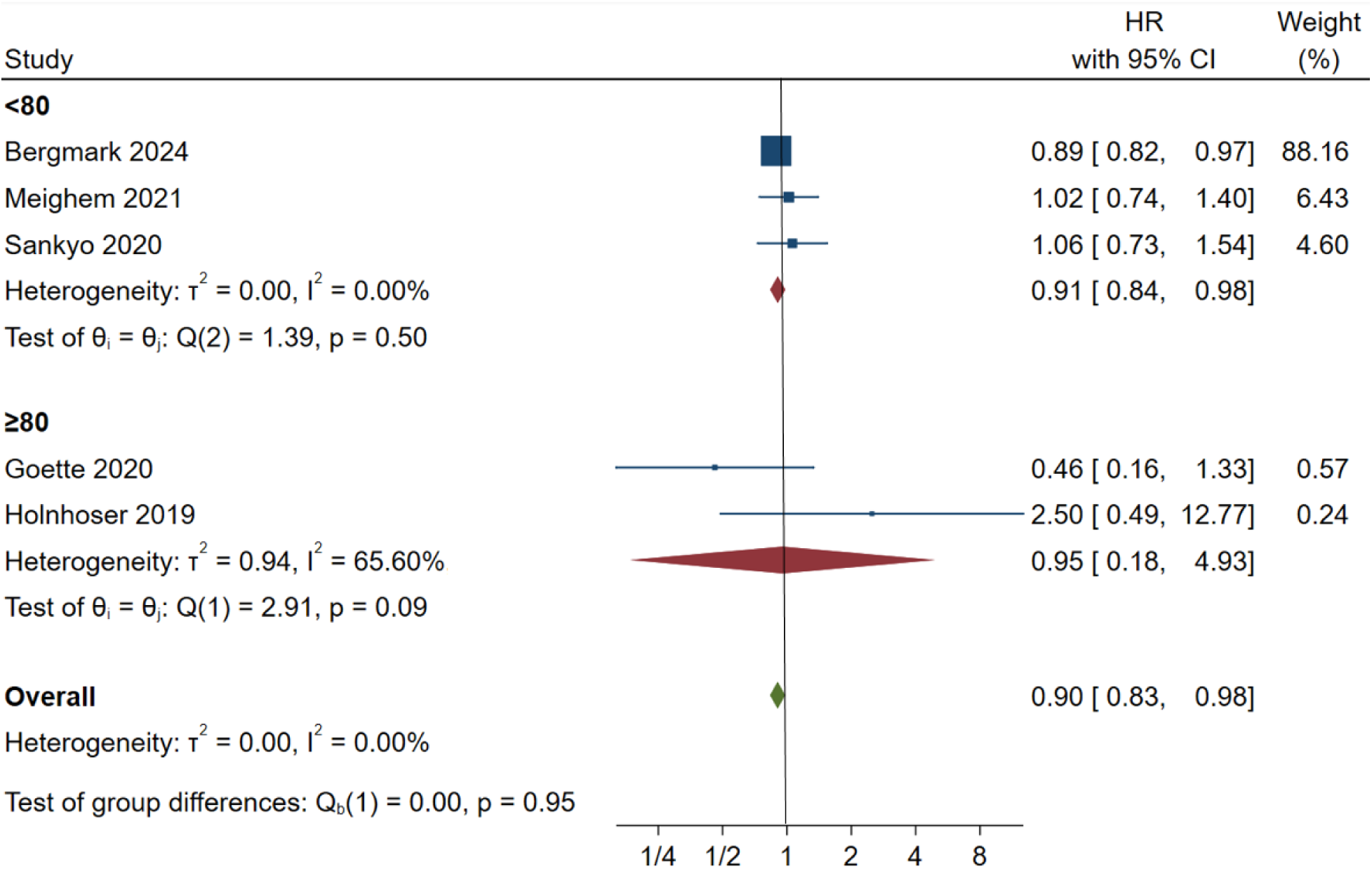
MACE CrCl

**Figure 3.**
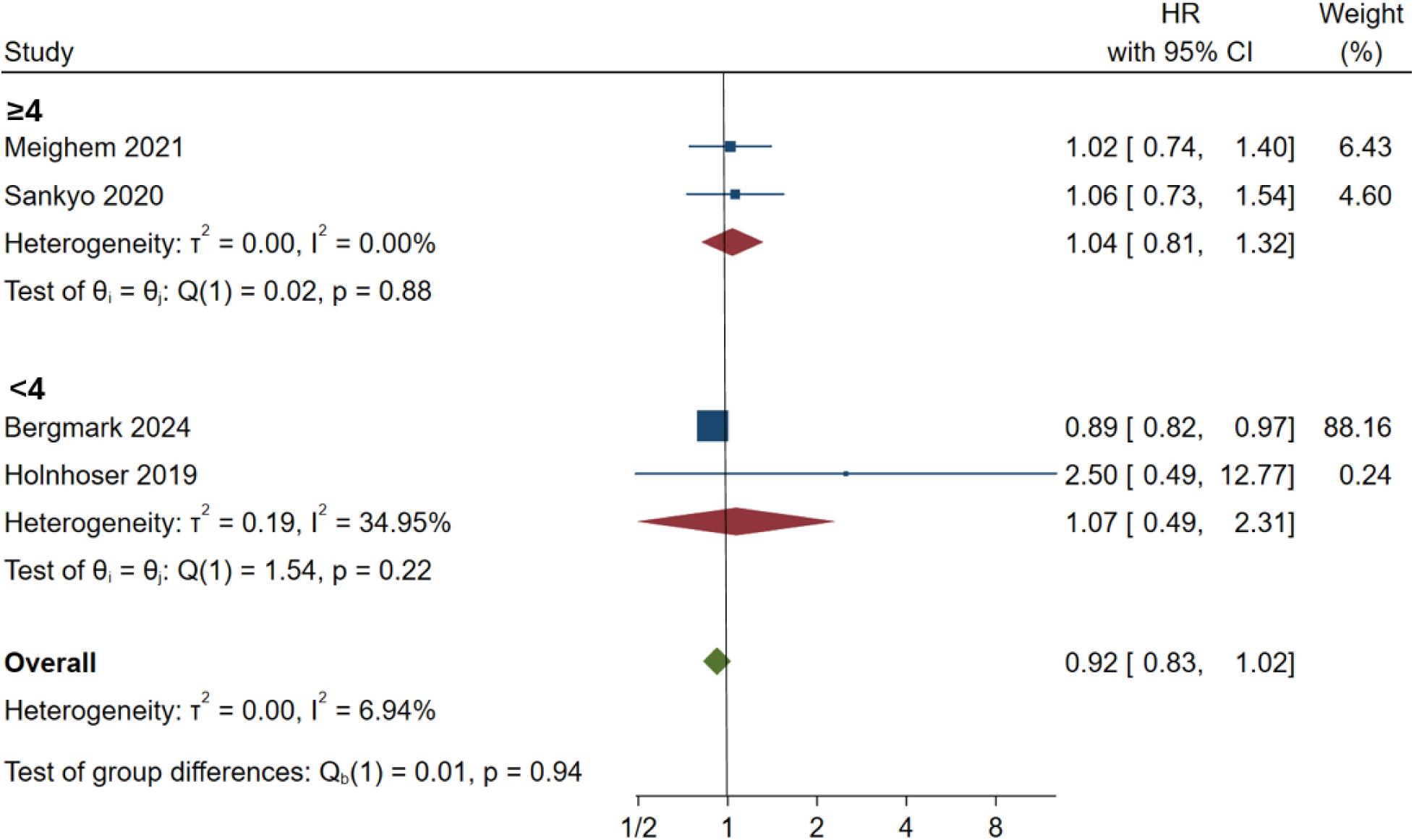
MACE CHA_2_DS_2_VASc

**Figure 4.**
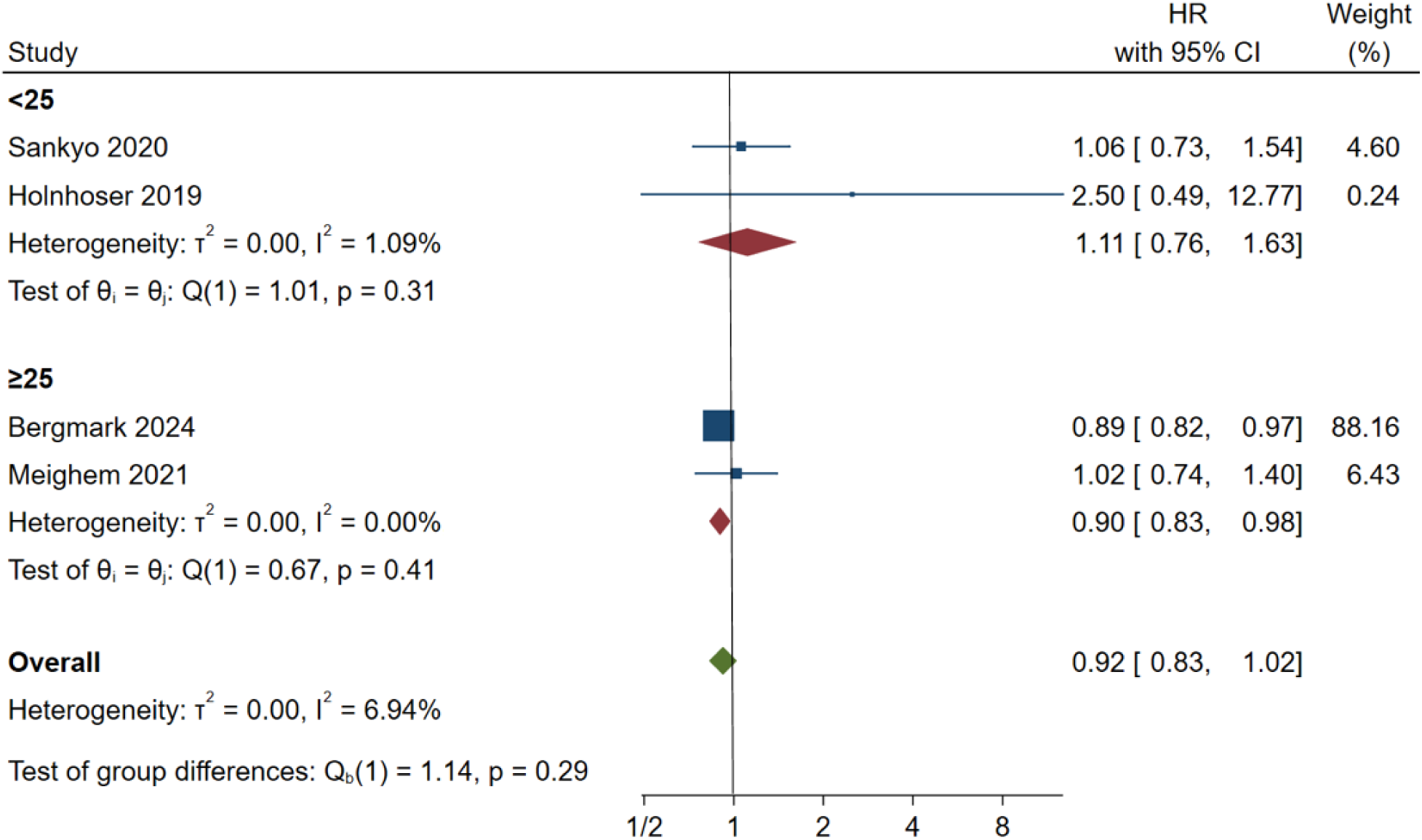
MACE heart failure

**Figure 5.**
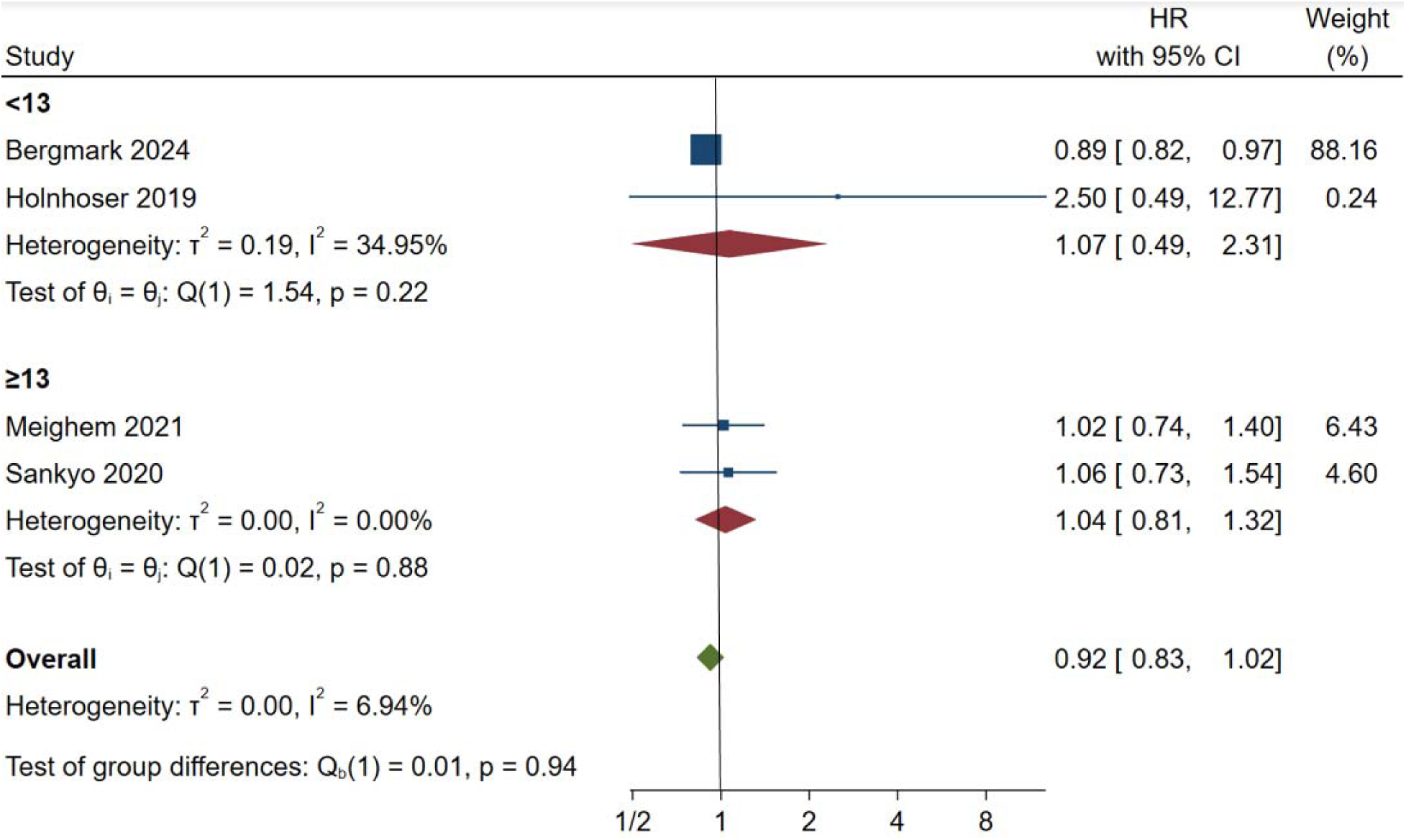
MACE previous MI

**Figure 6.**
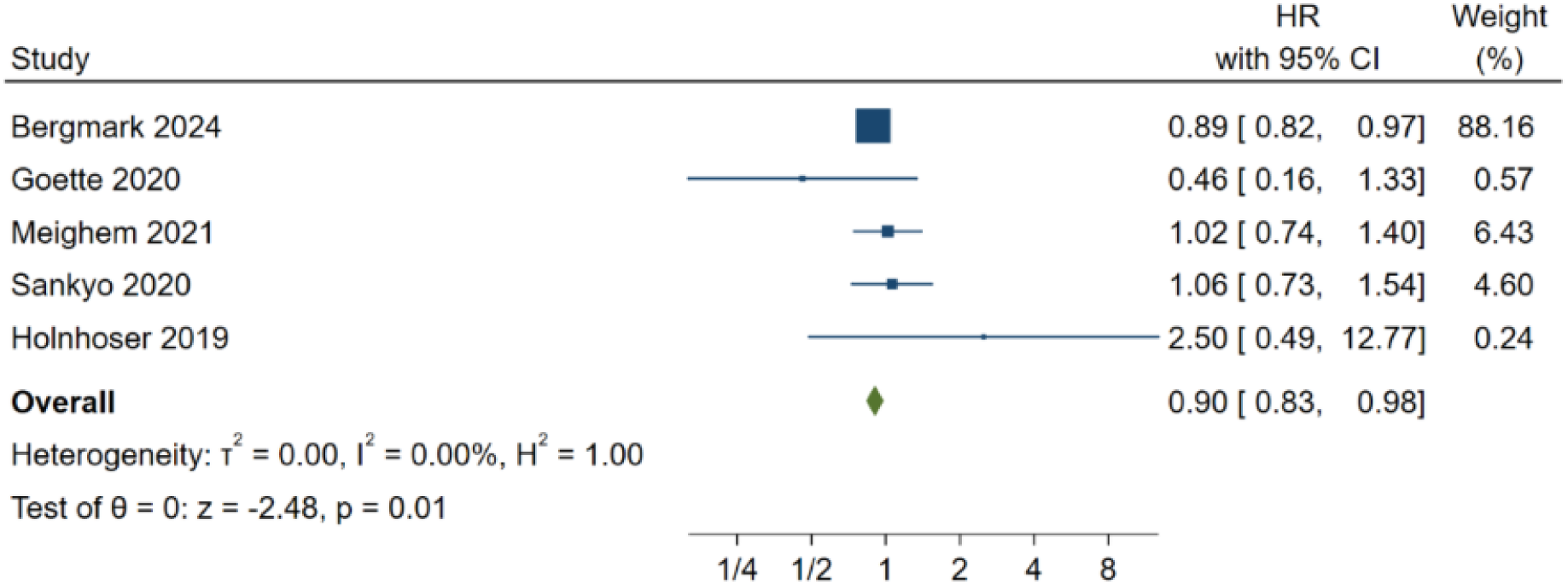
MACE age

**Figure 7.**
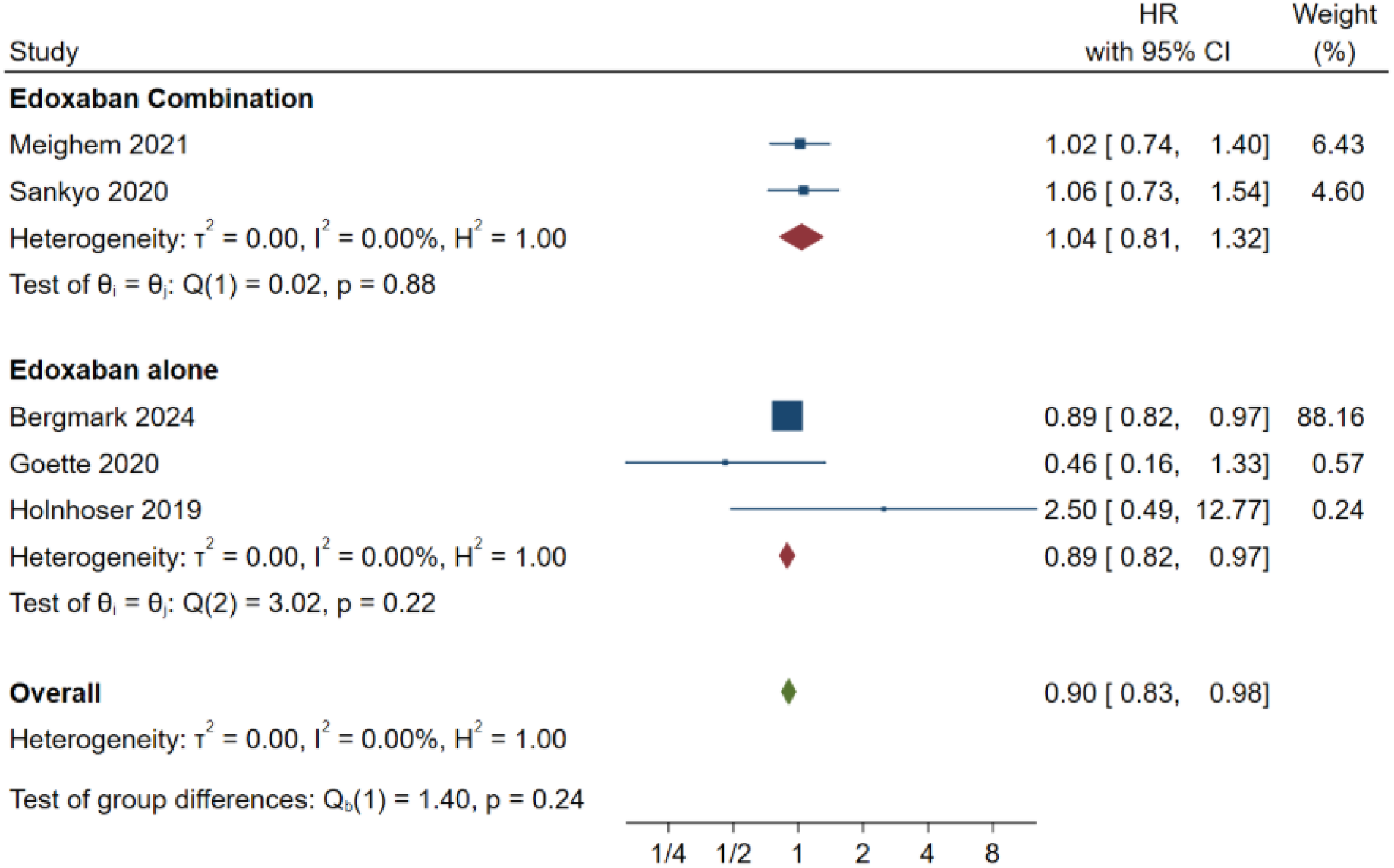
MACE with the type of therapy

### Stroke and Systemic Embolism (SSE)

Stroke and systemic embolism (SSE) outcomes analysis showed that edoxaban had a much greater effect in minimizing the probability of these events than warfarin with high heterogeneity (I2 = 92.2). This advantage was not, however, uniform in all groups of patients. The comparisons between the subgroups based on age and renal clearance provided also resulted in the statistically significant interactions (P < 0.05), indicating that the superiority of edoxaban in lessening the risk of SSE was rather constant across various demographic and clinical characteristics, particularly in younger patients and those with a higher creatinine clearance.

Edoxaban reduced the protective effect in patients with a history of a previous MI in the past (HR 0.58, 95% CI 0.32 - 1.05), suggesting that patients with a prior history of MI do not get the same amount of risk reduction in SSE as the general population. [Figures 8-13]

**Figure 8.**
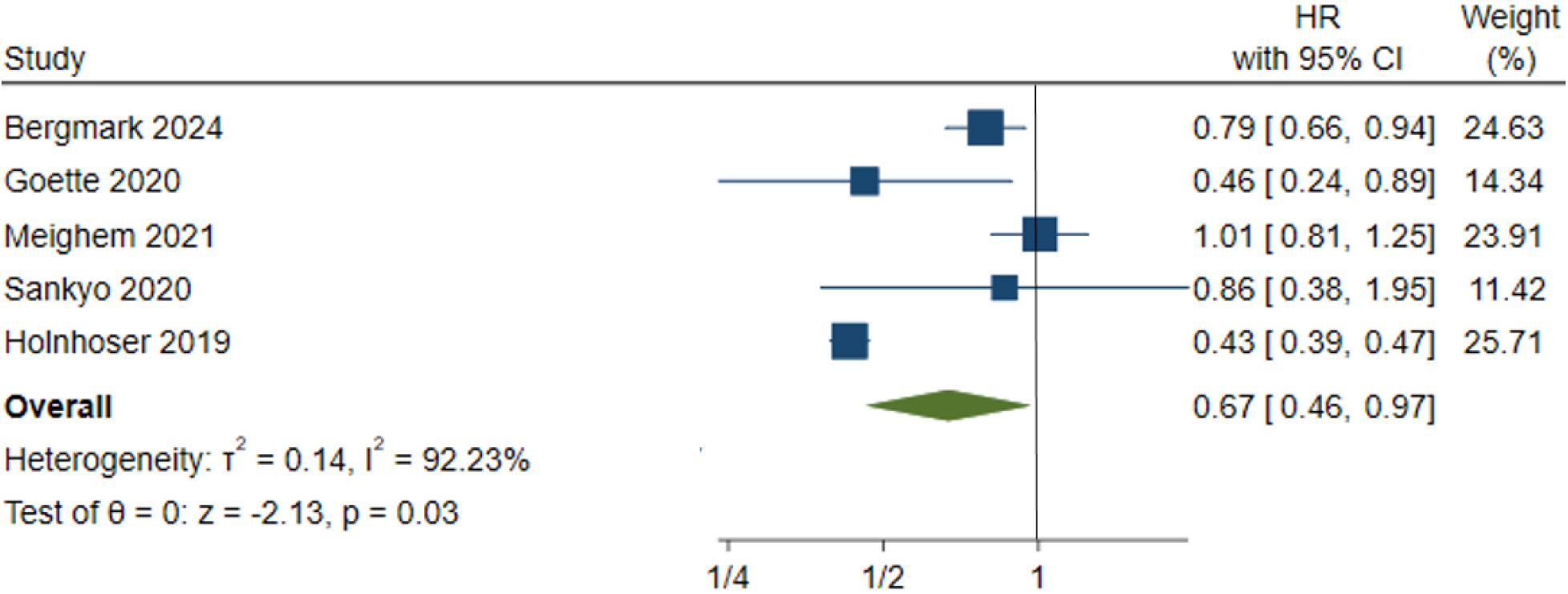
Stroke and systemic embolism

**Figure 9.**
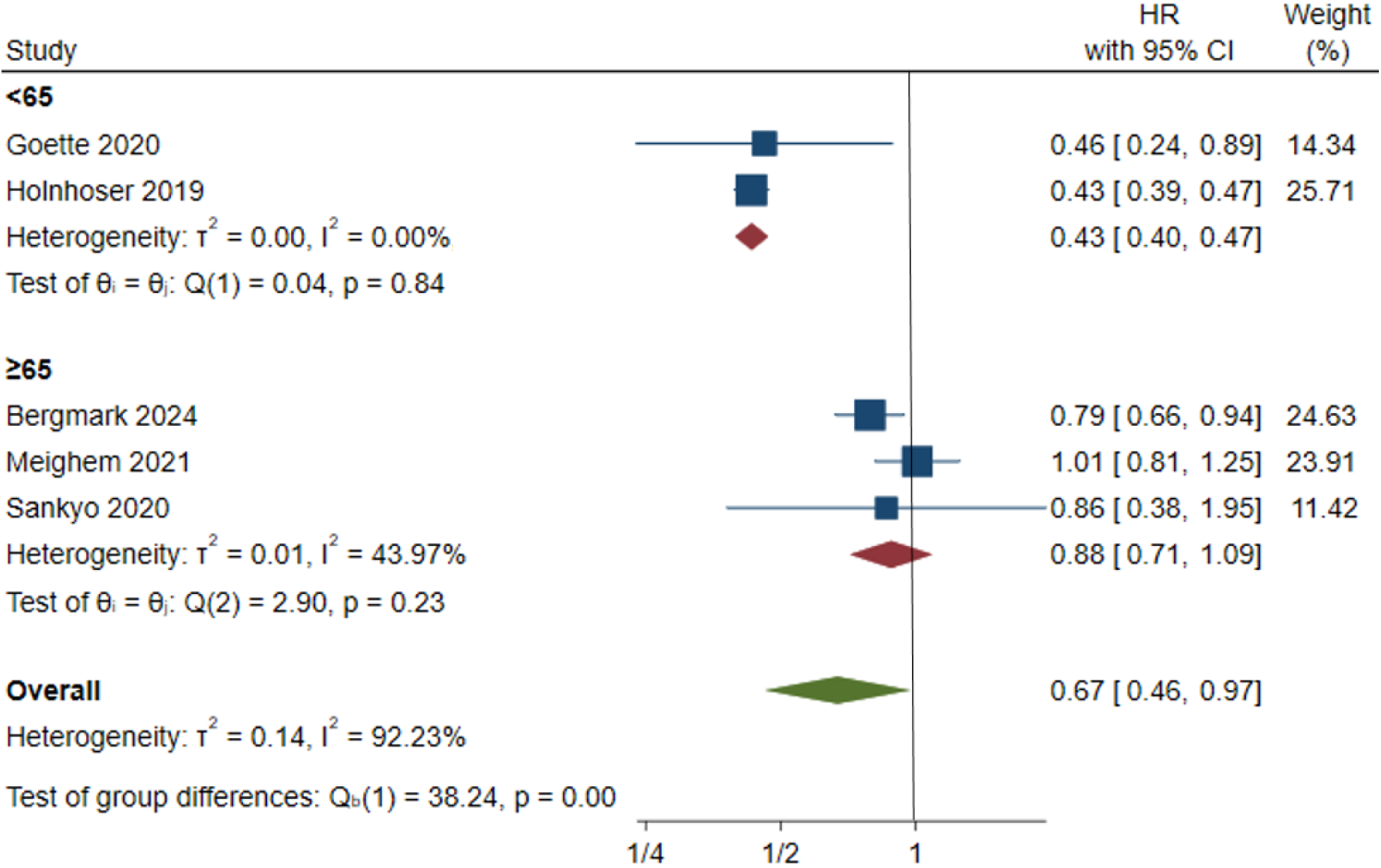
SSE age

**Figure 10.**
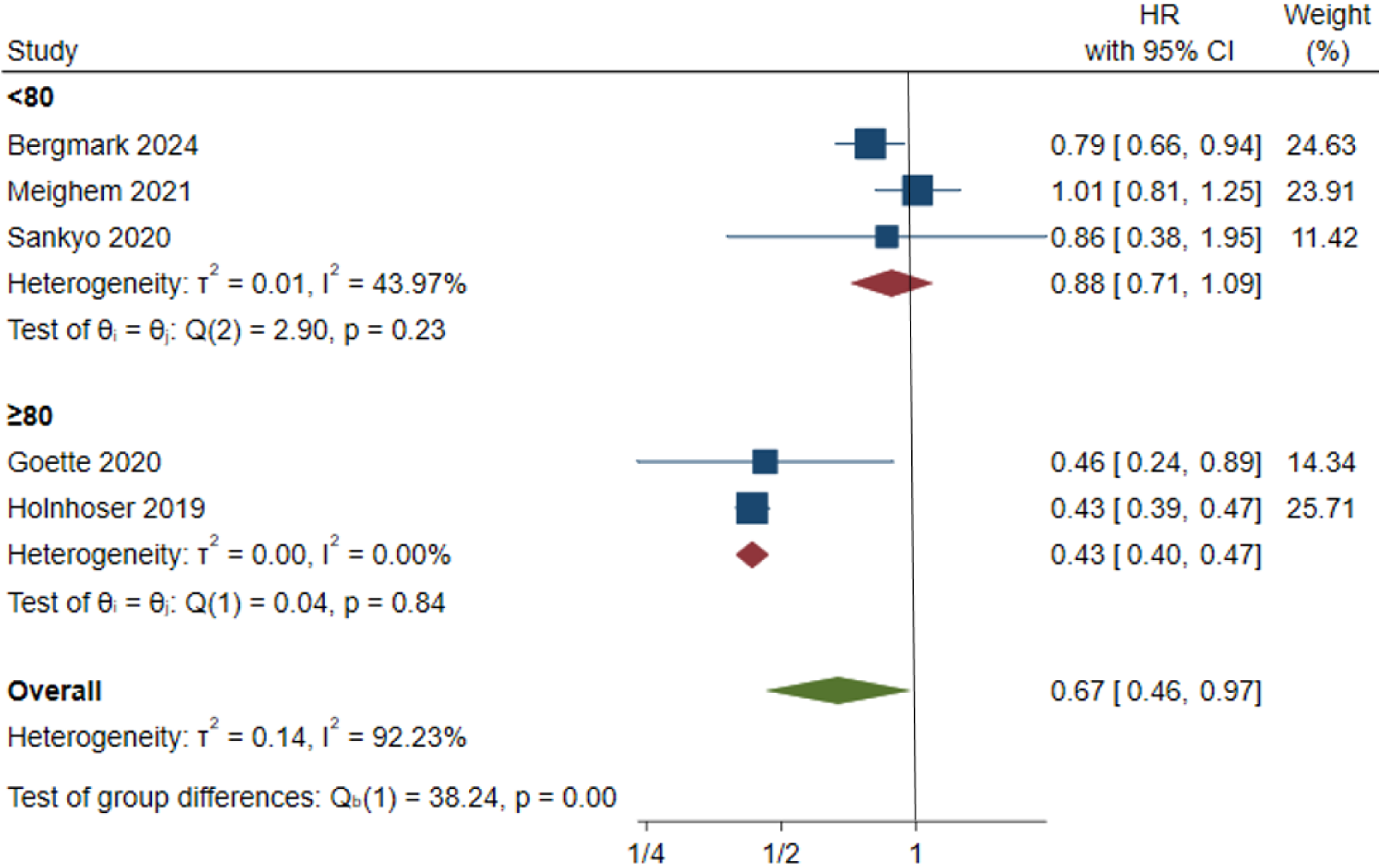
SSE CrCl

**Figure 11.**
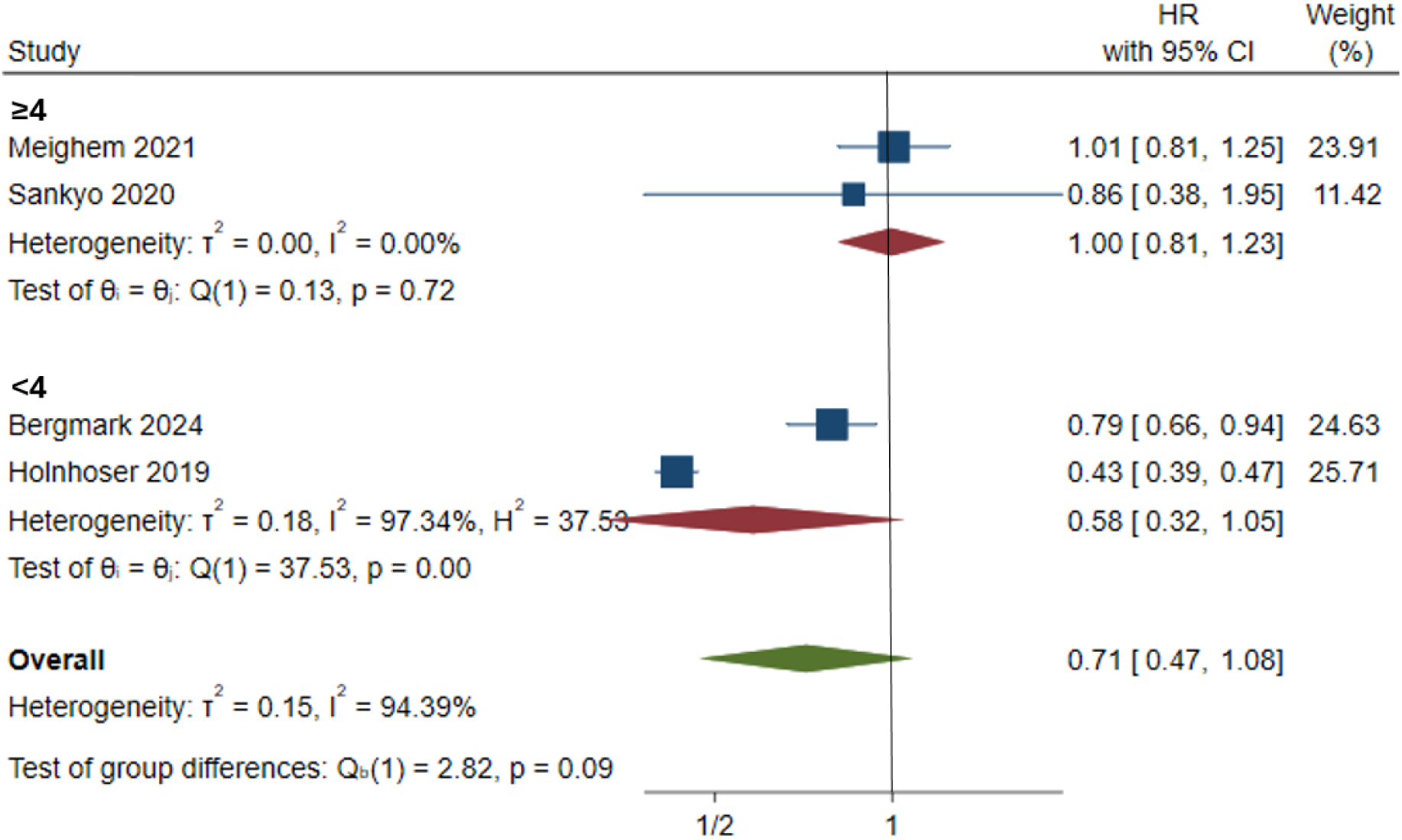
SSE HA_2_DS_2_VASc

**Figure 12.**
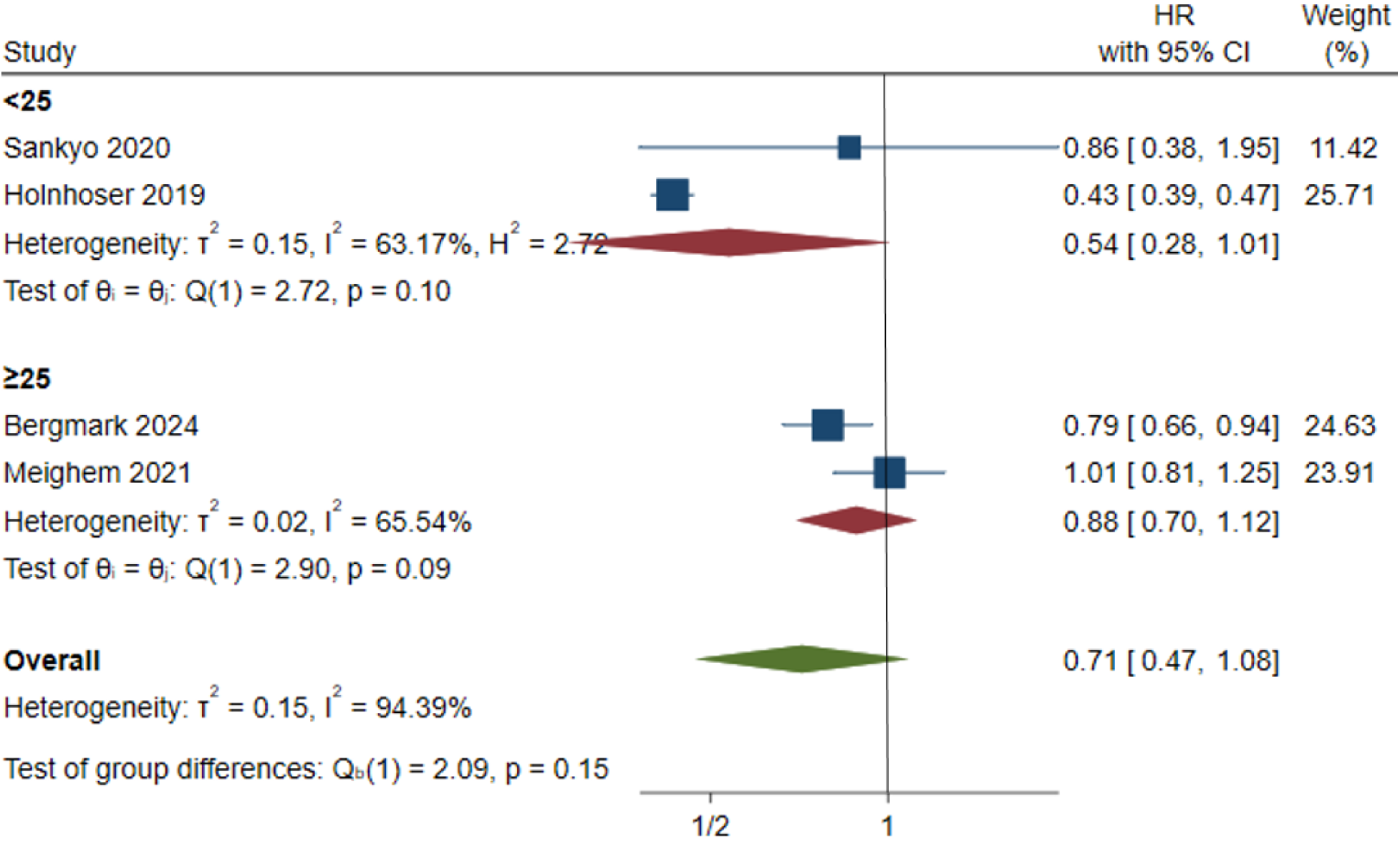
SSE heart failure

**Figure 13.**
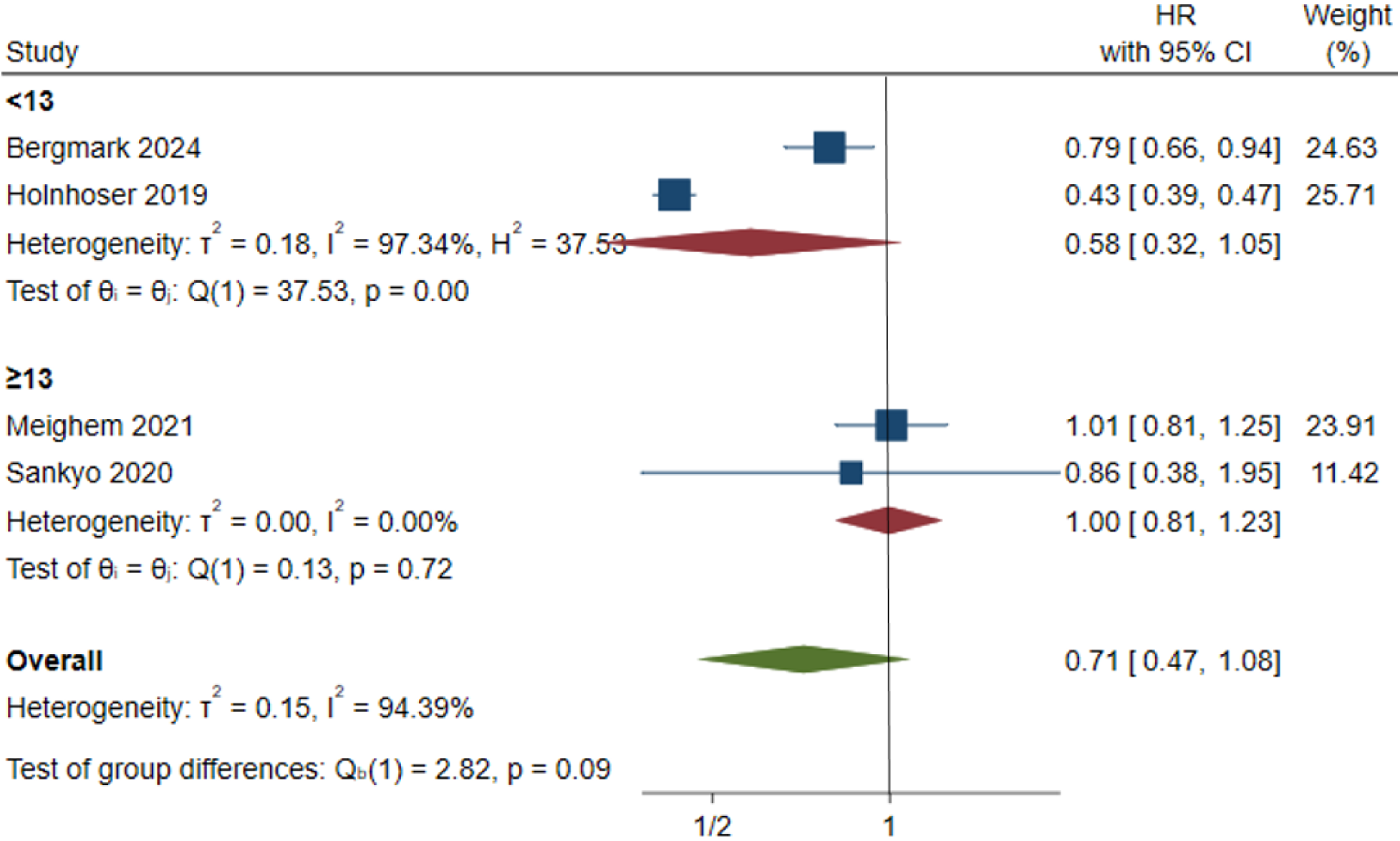
SSE previous MI

### All-cause mortality (ACM)

In the analysis of all-cause mortality, there were no statistically significant differences between edoxaban and warfarin (HR = 0.65, p > 0.05), which proved that agents did not show a definite survival benefit compared to each other. This observation was similar in the different subgroups, which were stratified by age, renal of heart failure status, and renal functioning. The absence of difference in all-cause mortality indicates that whereas the use of edoxaban might prove beneficial in some cardiovascular and bleeding-related consequences, the same does not always lead to better survival as observed in Figures 14-16.

**Figure 14.**
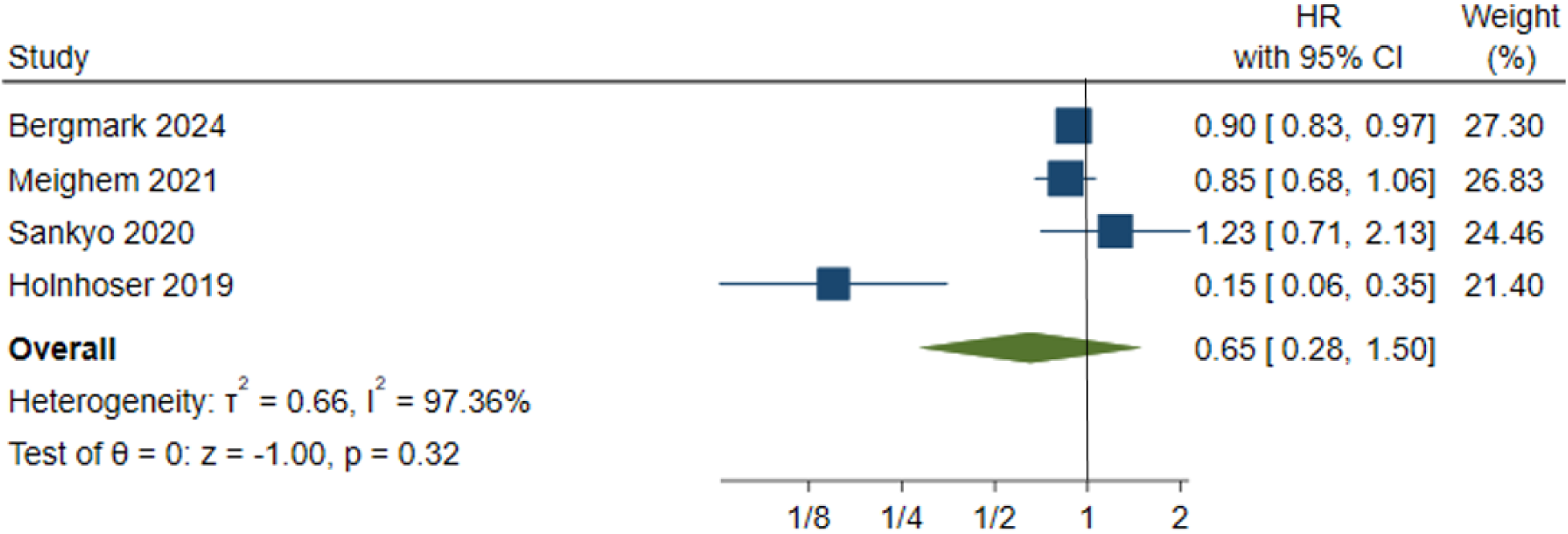
All-cause mortality

**Figure 15.**
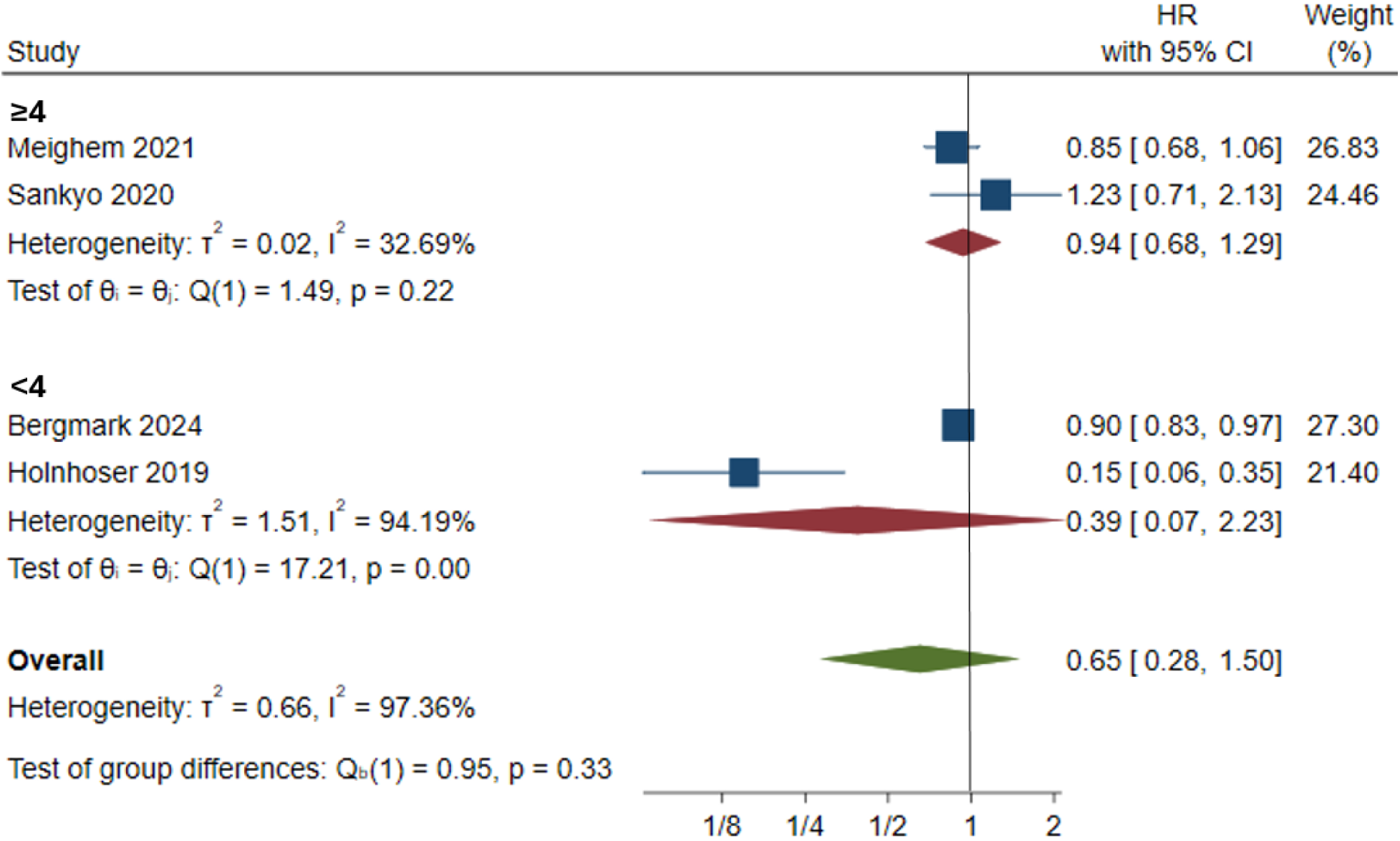
ACM CHA_2_DS_2_VASc

**Figure 16.**
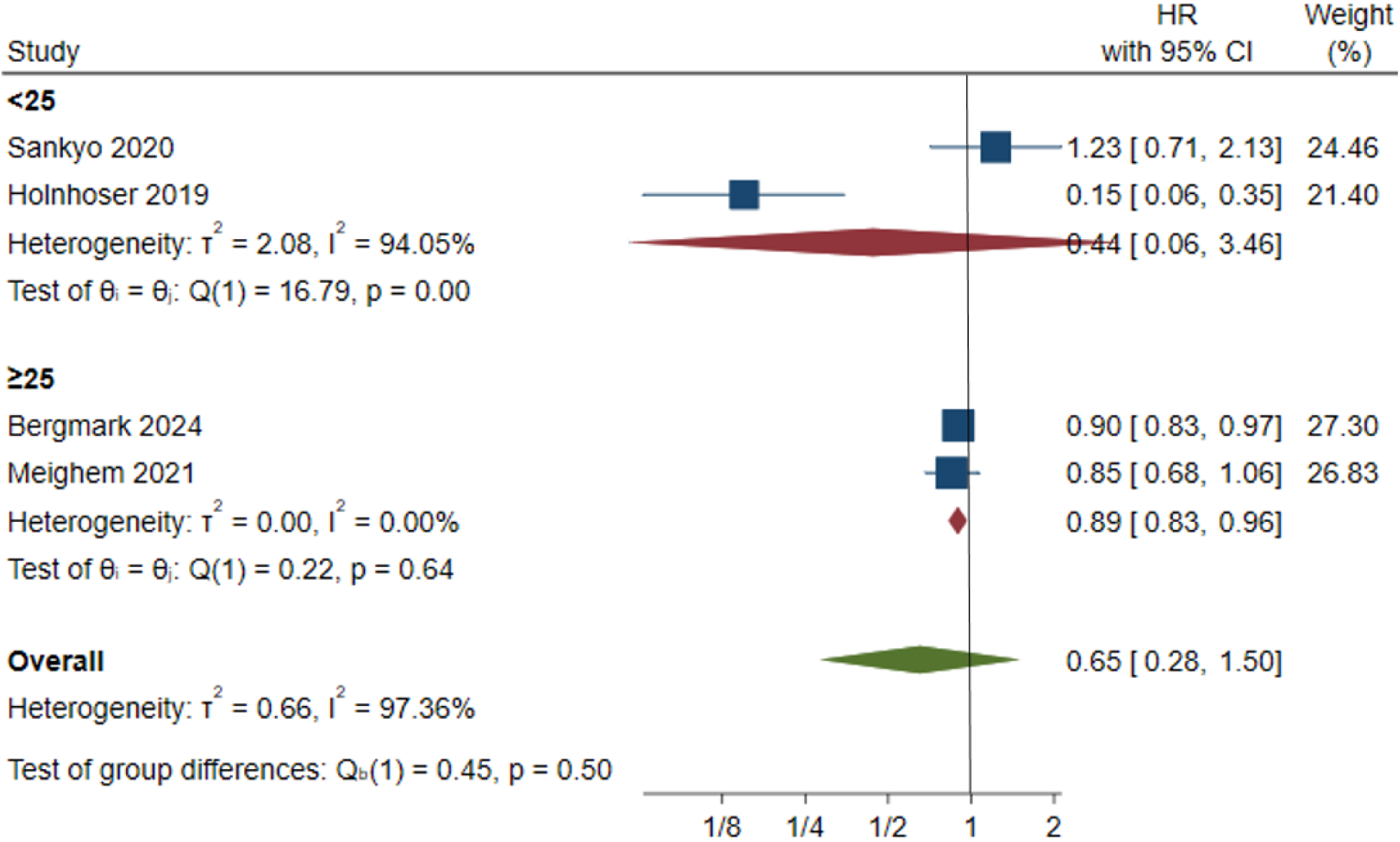
ACM heart failure

### Major bleeding (MB)

Risk of significant bleeding was not any different between edoxaban and warfarin with a hazard ratio of 0.95 (95% CI 0.60-1.50, p = 0.83). This result supports the idea that edoxaban can be the alternative with a lower risk of bleeding. Nevertheless, stratification by the burden of heart failure has shown that the effect of edoxaban was greater in those populations with the prevalence of HF less than 25. Conversely in populations where HF prevalence was >=25 warfarin seemed to confer a minor and consequently non-significant benefit in the risk of major bleeding (HR 0.95, I2 = 93.0). Aged patients above the age of 65 demonstrated this with edoxaban being better than warfarin compared to younger patients where the vice versa was observed. Significant differences were not found within other clinical characteristics CrCl, CHA2DS2VASc and previous MI. This was shown in figures 17-22.

**Figure 17.**
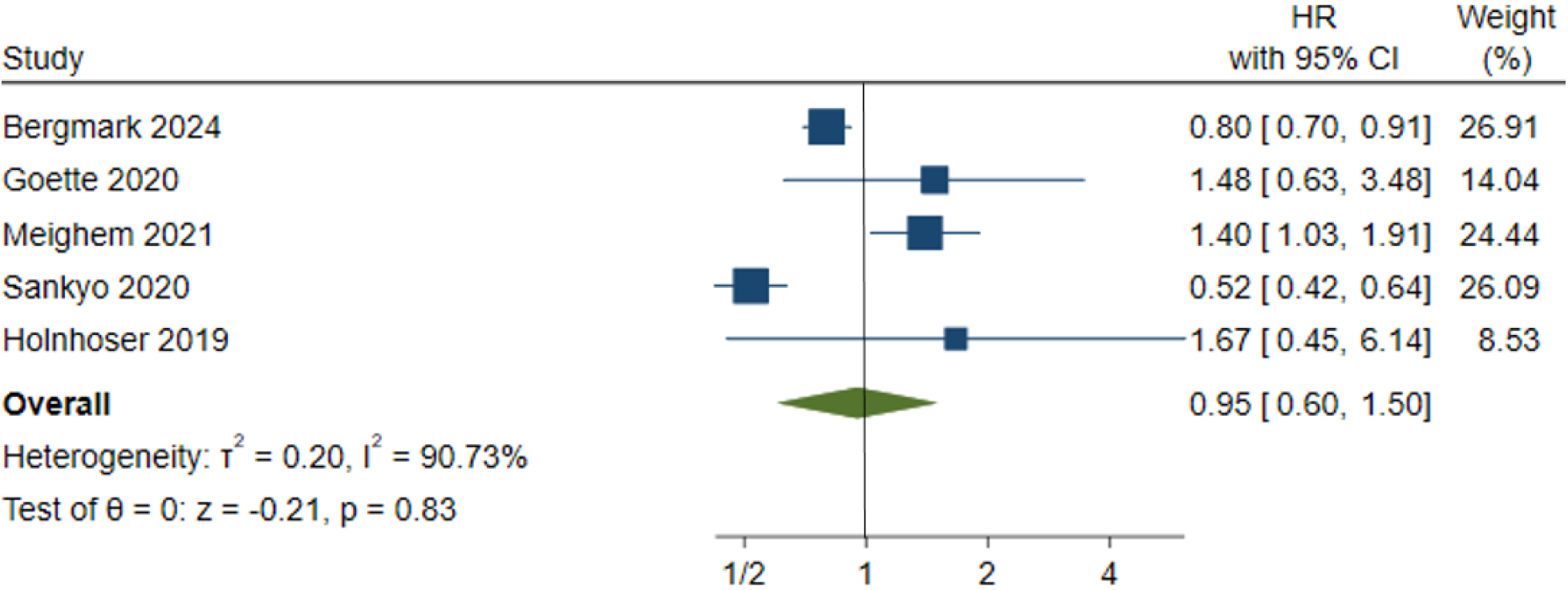
Major bleeding

**Figure 18.**
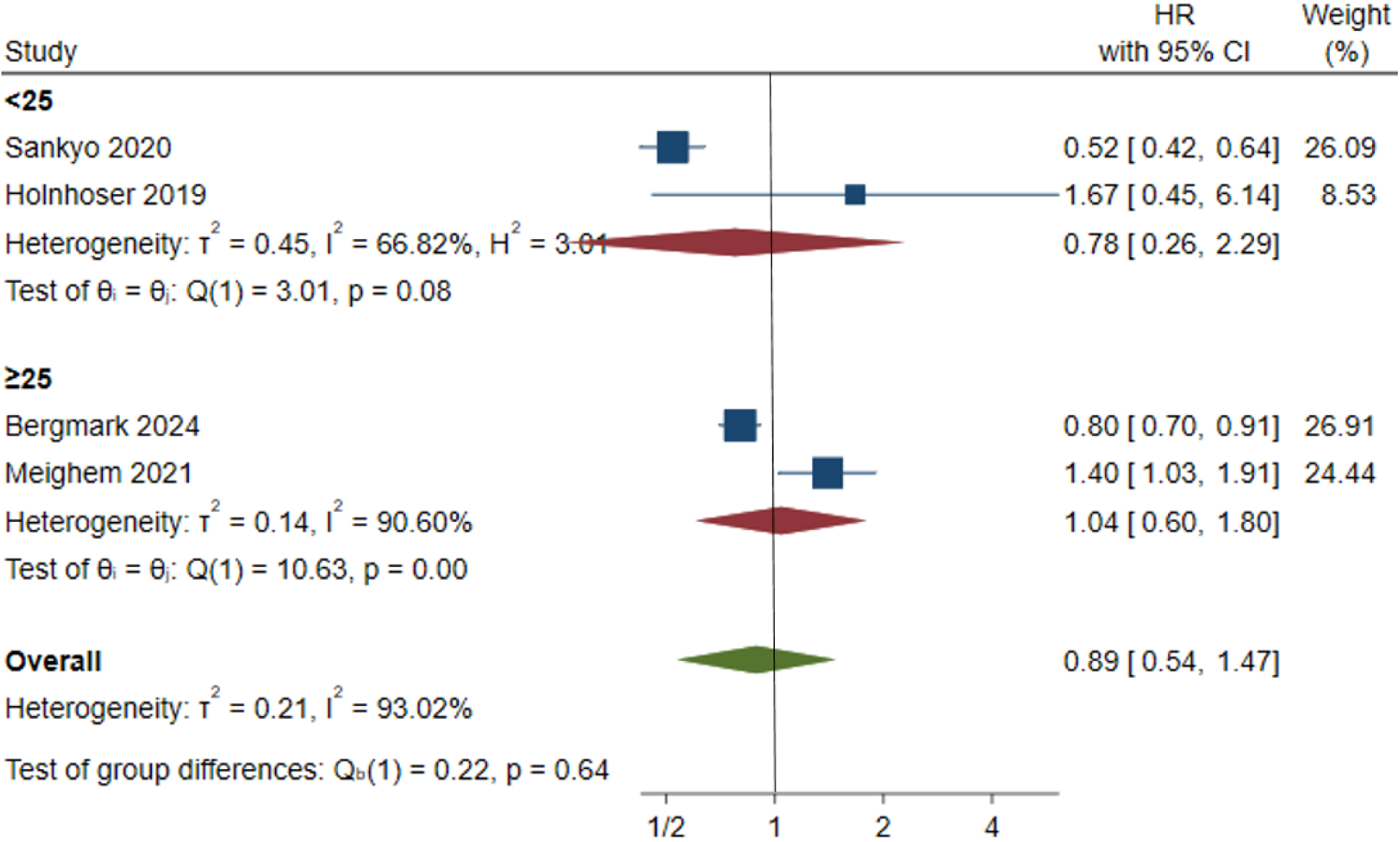
MB heart failure

**Figure 19.**
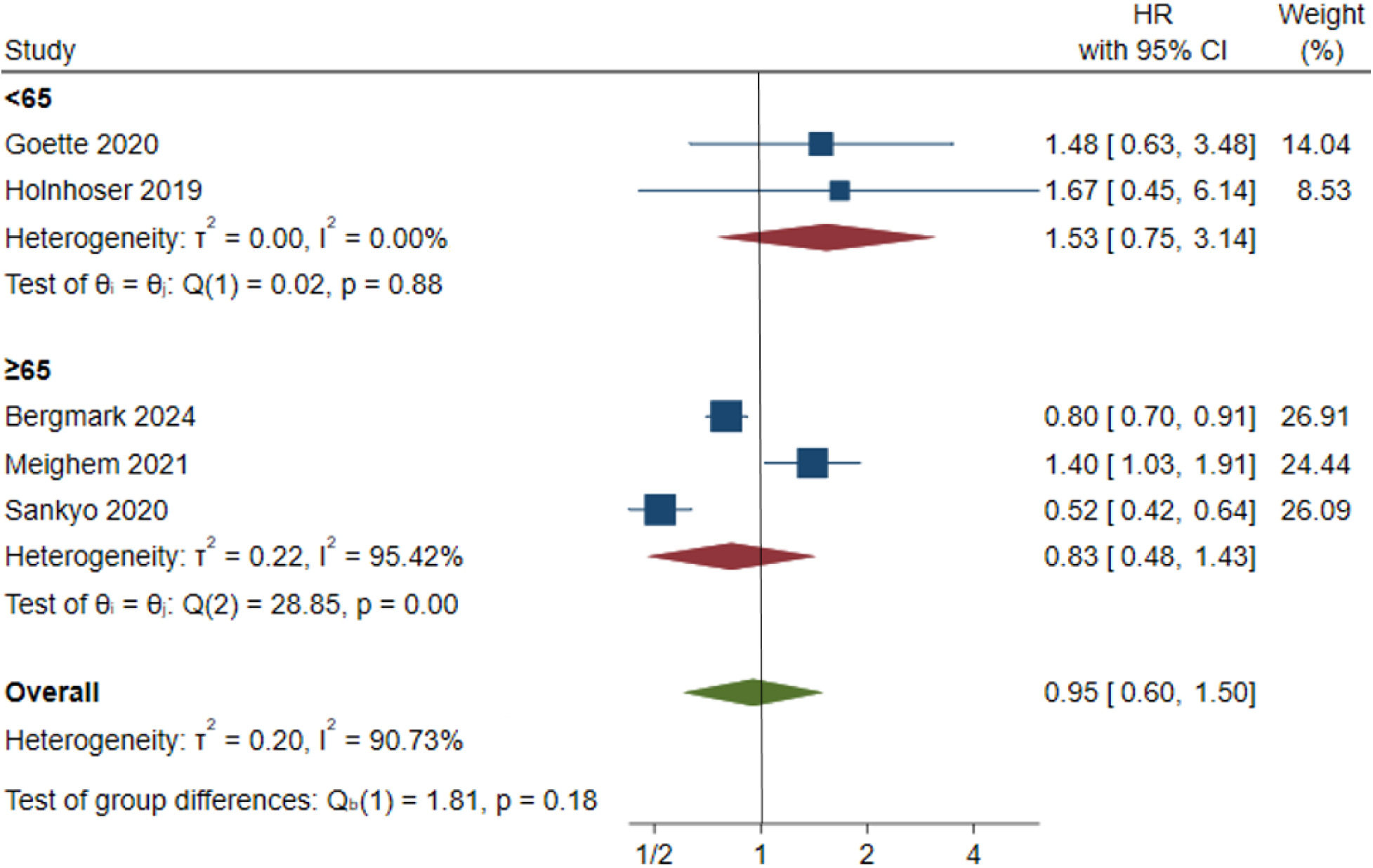
MB age

**Figure 20.**
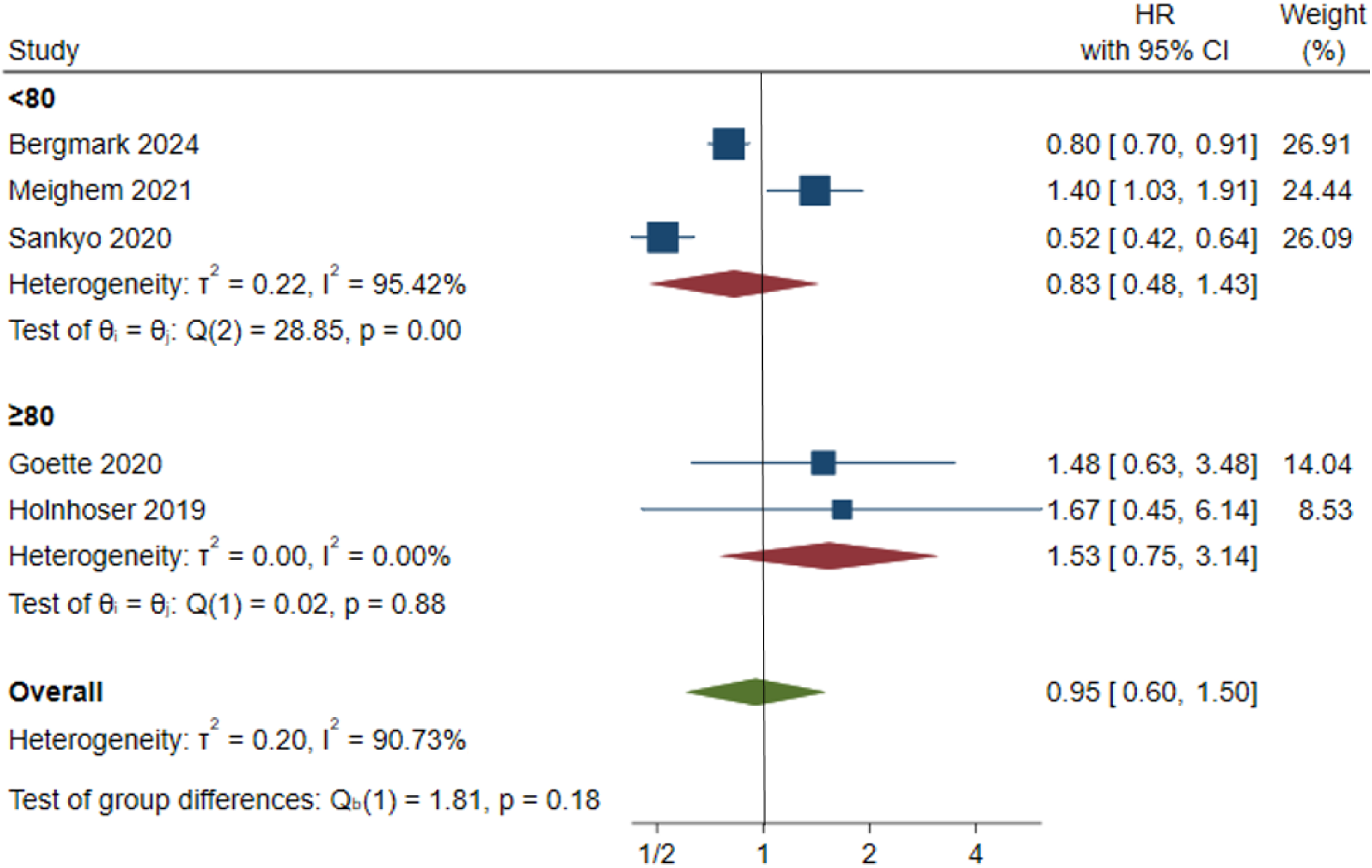
MB CrCl

**Figure 21.**
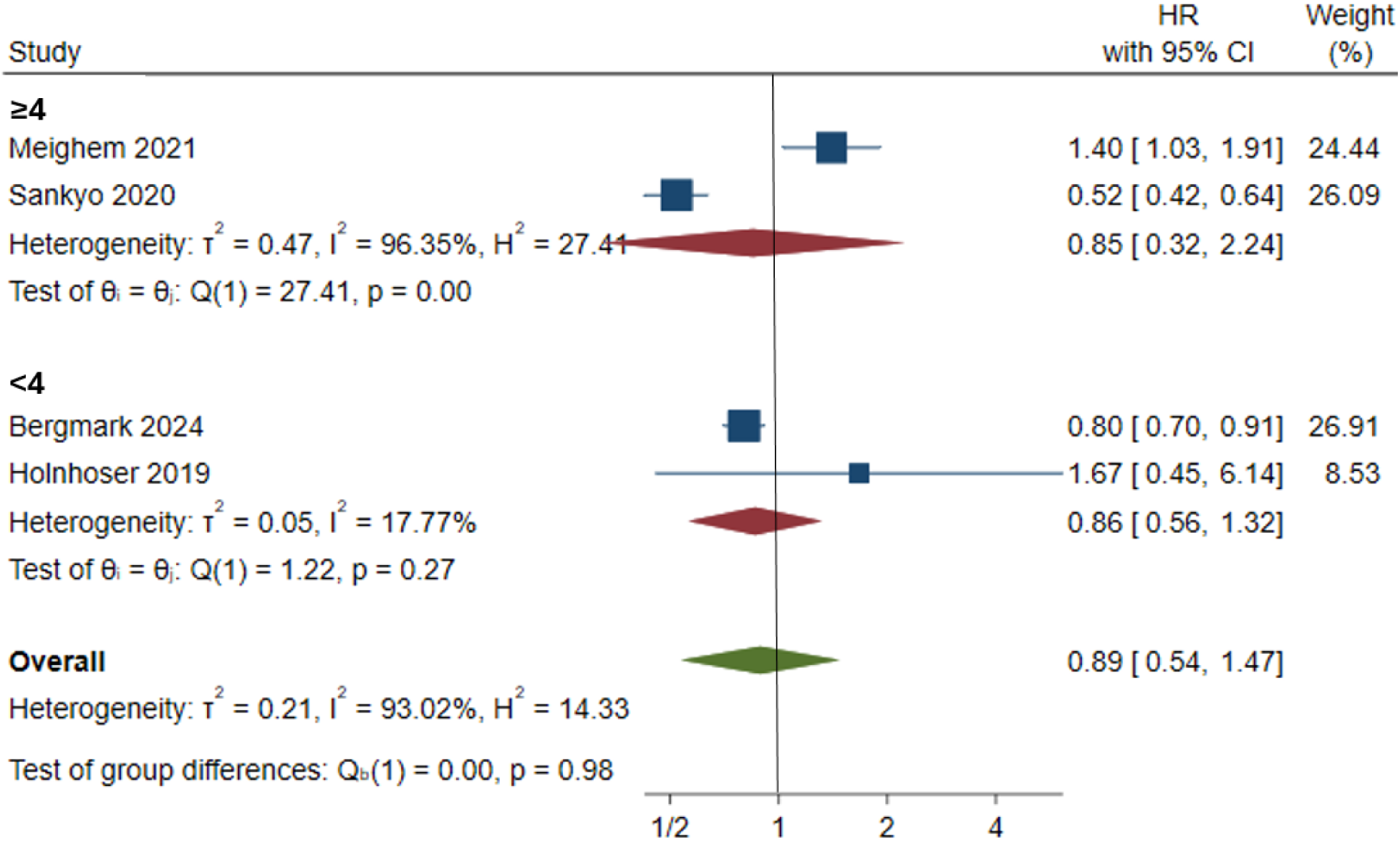
MB CHA_2_DS_2_VASc

**Figure 22.**
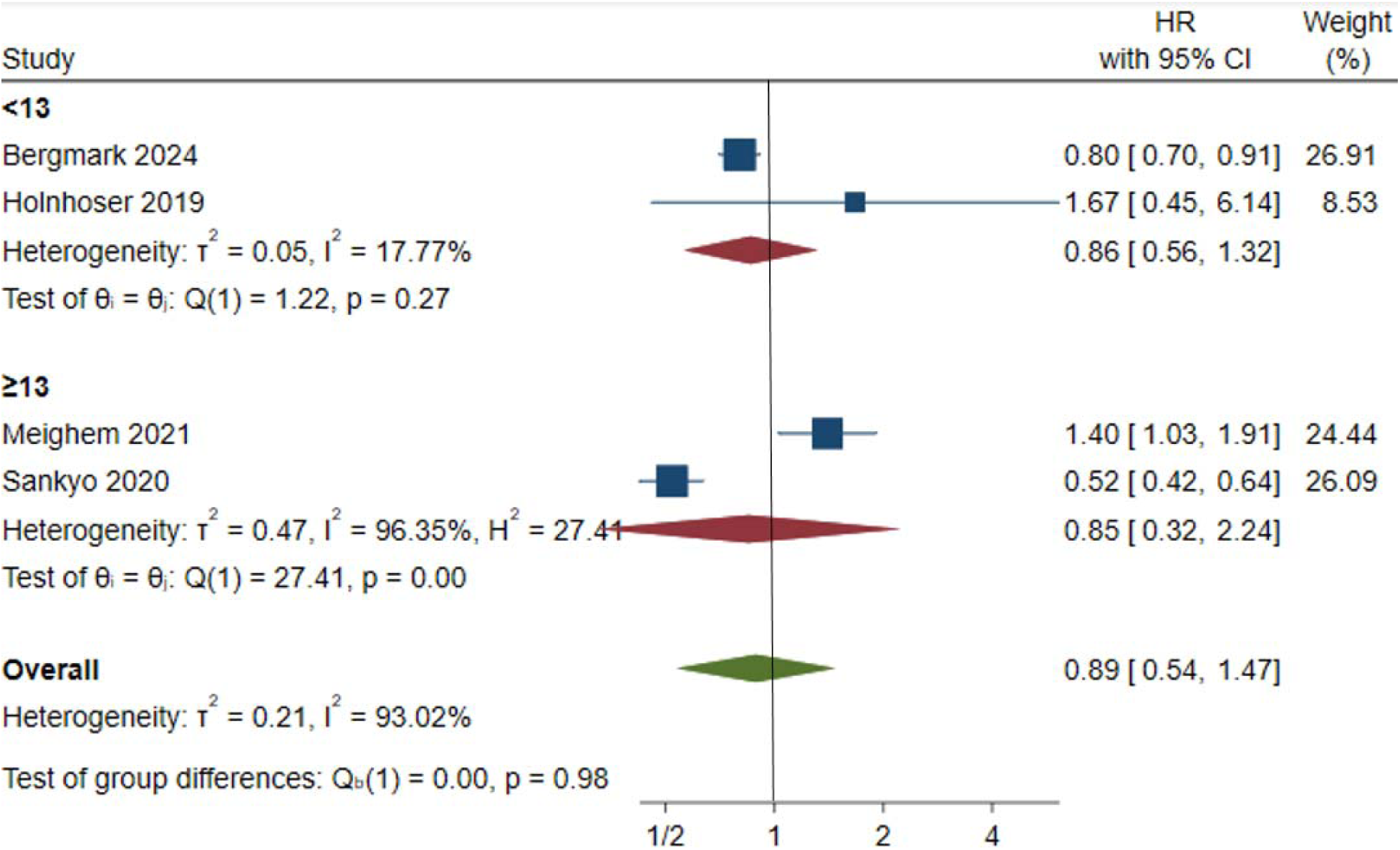
MB previous MI

### Clinically relevant non-major bleeding (CRNM)

Edoxaban and warfarin had a similar risk of clinically relevant non-major bleeding (CRNM) with a hazard ratio of (1.19, 95% CI 0.75 - 1.91, p = 0.46). The warfarin in younger patients (under 65 years) was linked with a greater CRNM risk but not significantly different (HR 2.49, p = 0.90). The heterogeneity was reported to be high. These variations underscore the need to customize the anticoagulant treatment of patients in accordance with the specifics of patients. Additional subgroup analyses based on CHA2DS2-VASc-score and previous MI-status further underscored the complexity of bleeding risk prediction in which warfarin has been found to have better effects in high-risk groups, whereas edoxaban is still a better choice in patients with lower thromboembolic risk profiles.

**Figure 23.**
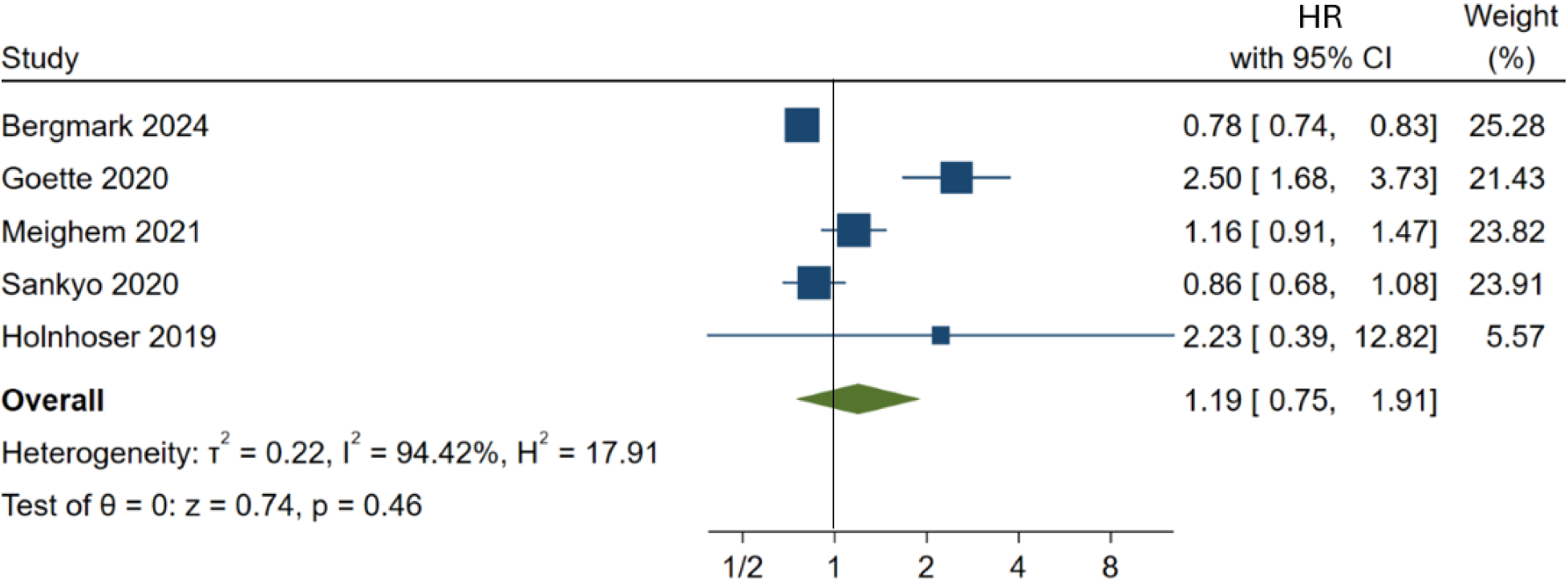
Clinically relevant non-major bleeding

**Figure 24.**
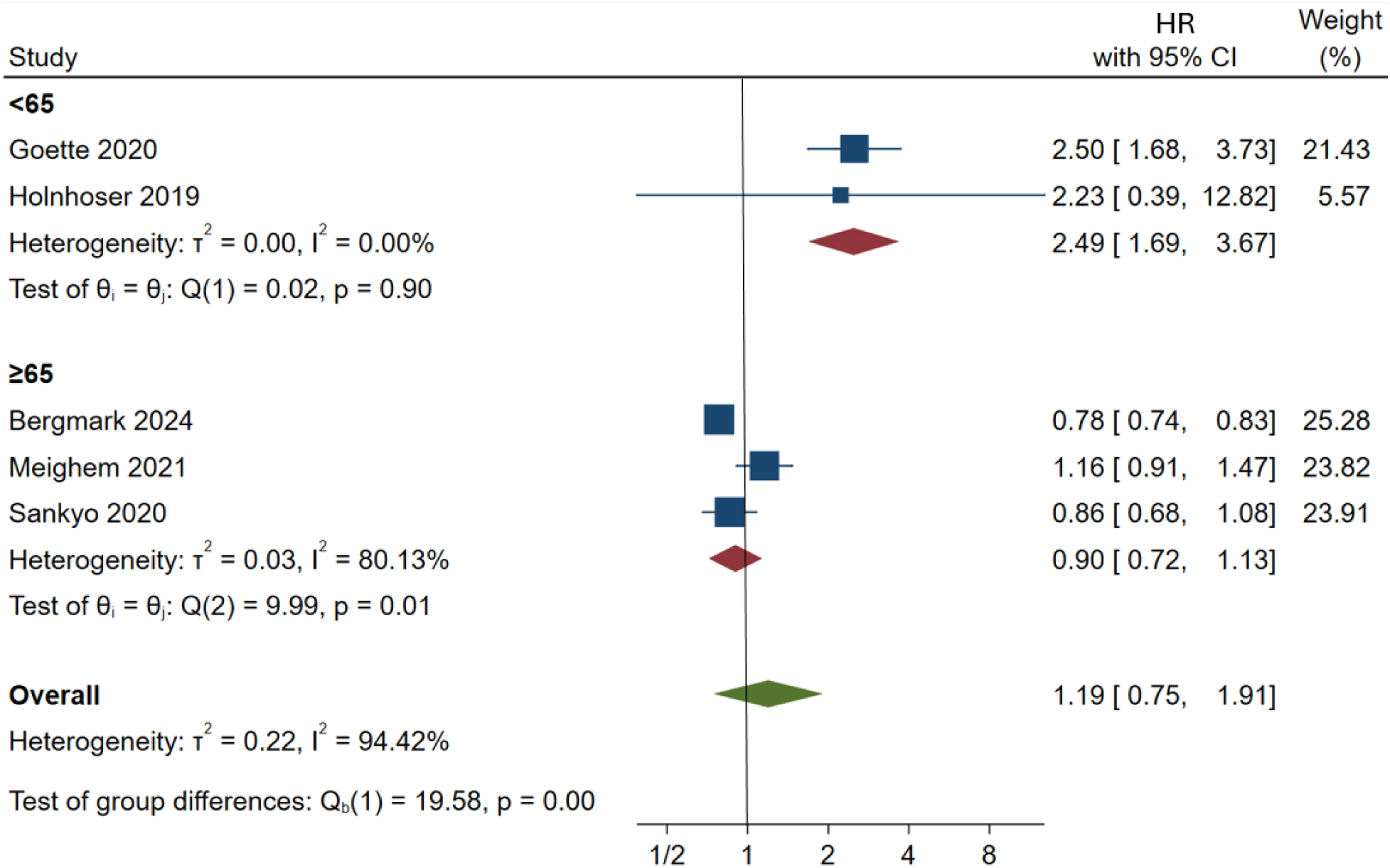
CRNM age

**Figure 25.**
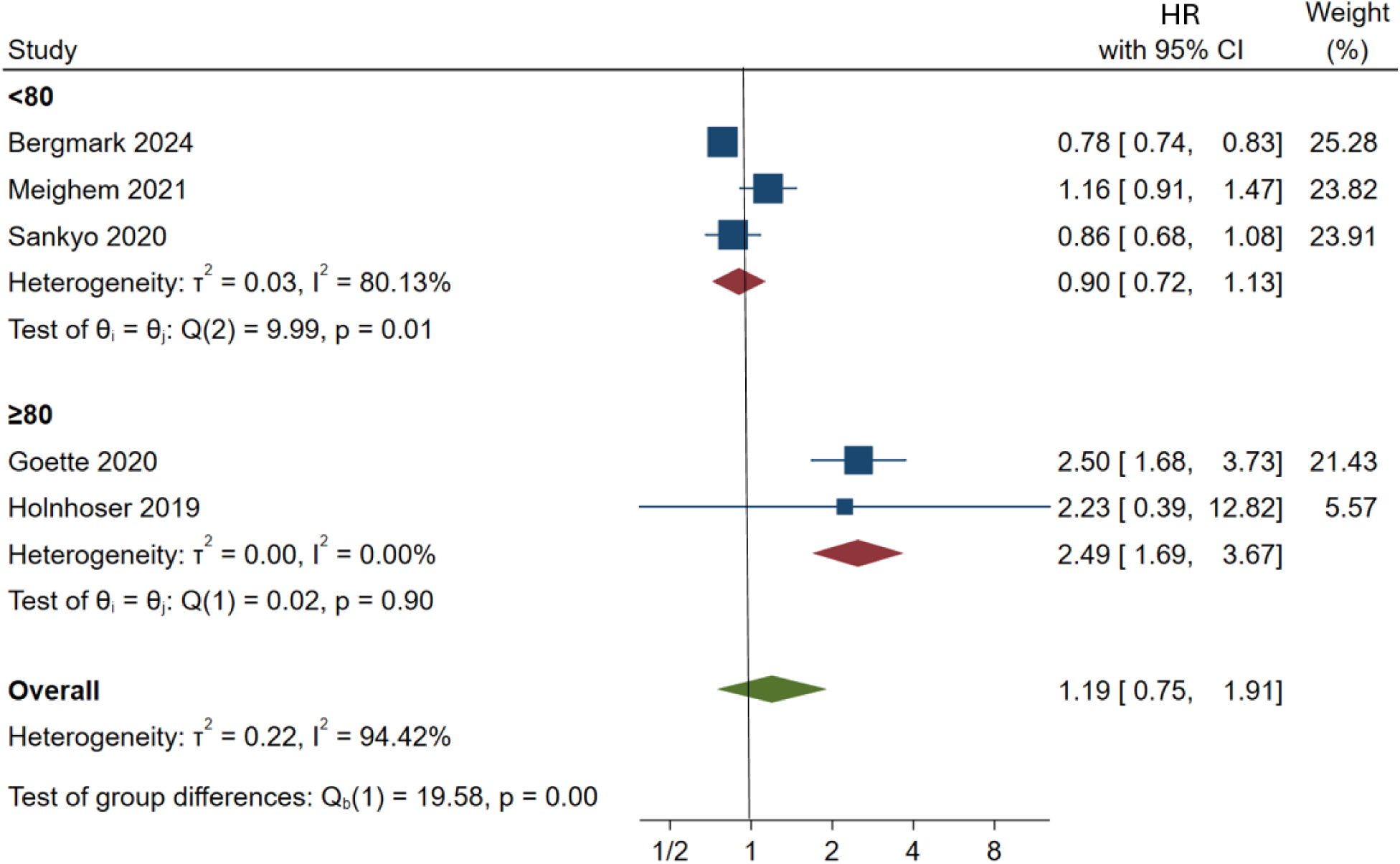
CRNM CrCl

**Figure 26.**
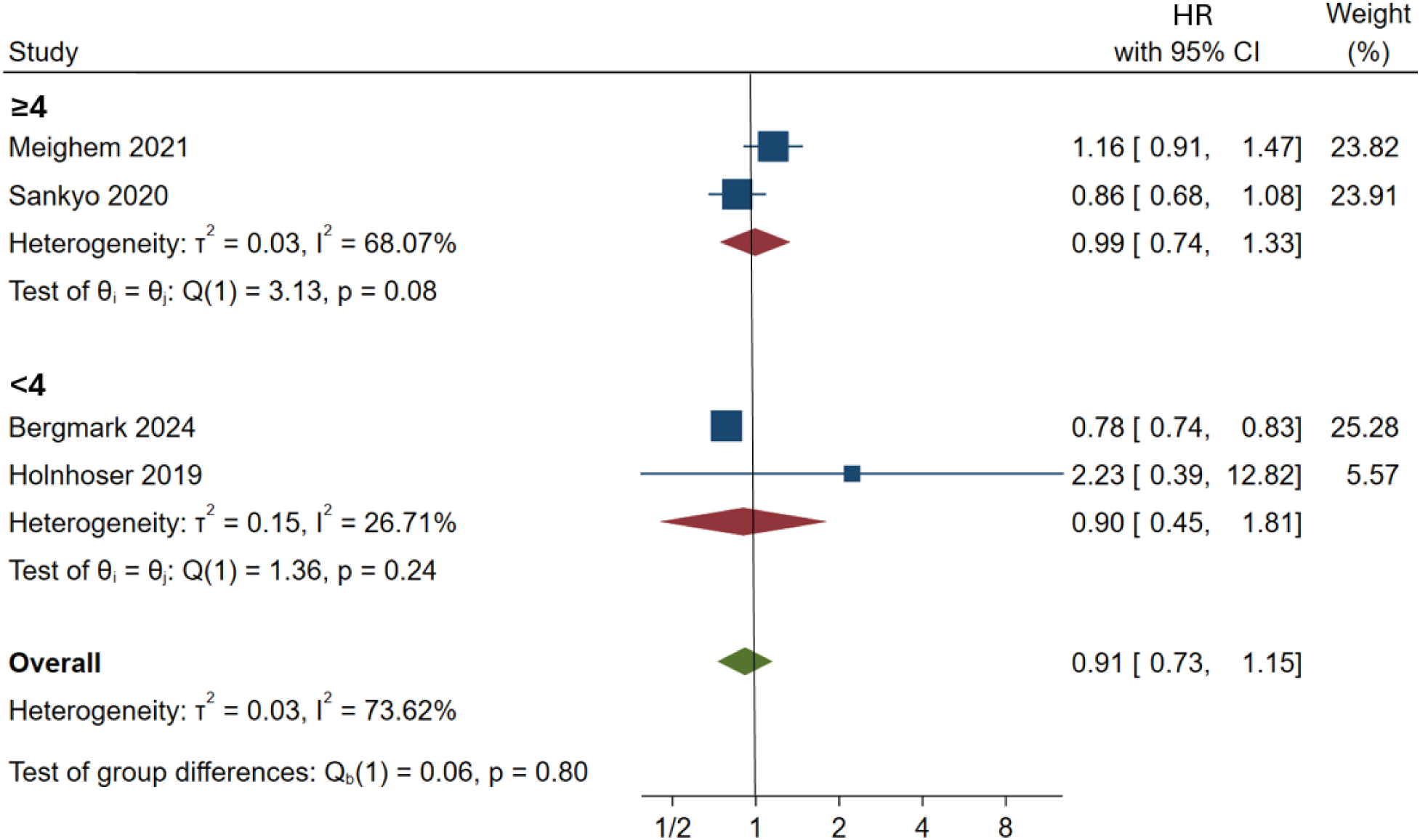
CRNM CHA_2_DS_2_VASc

**Figure 27.**
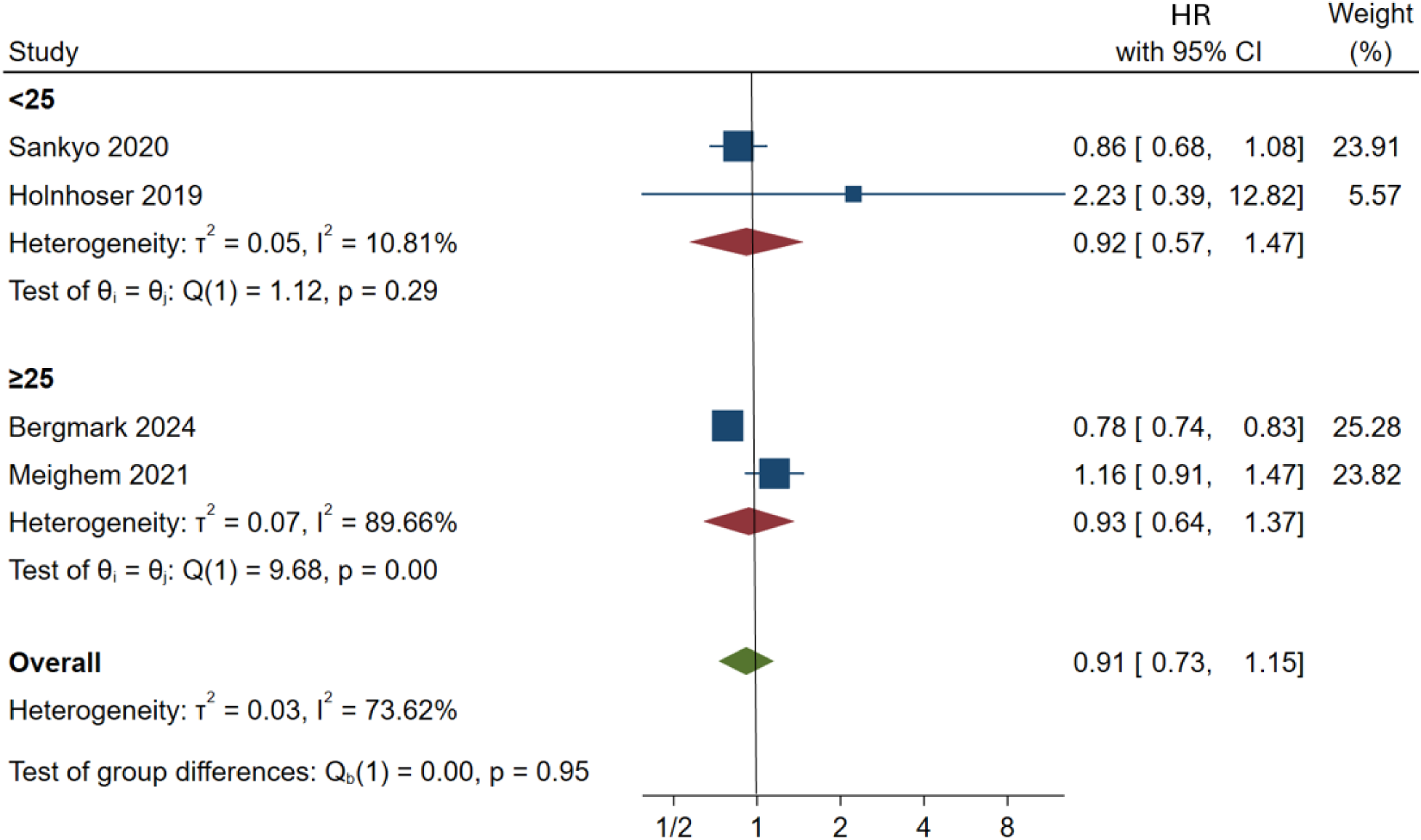
CRNM HF

**Figure 28.**
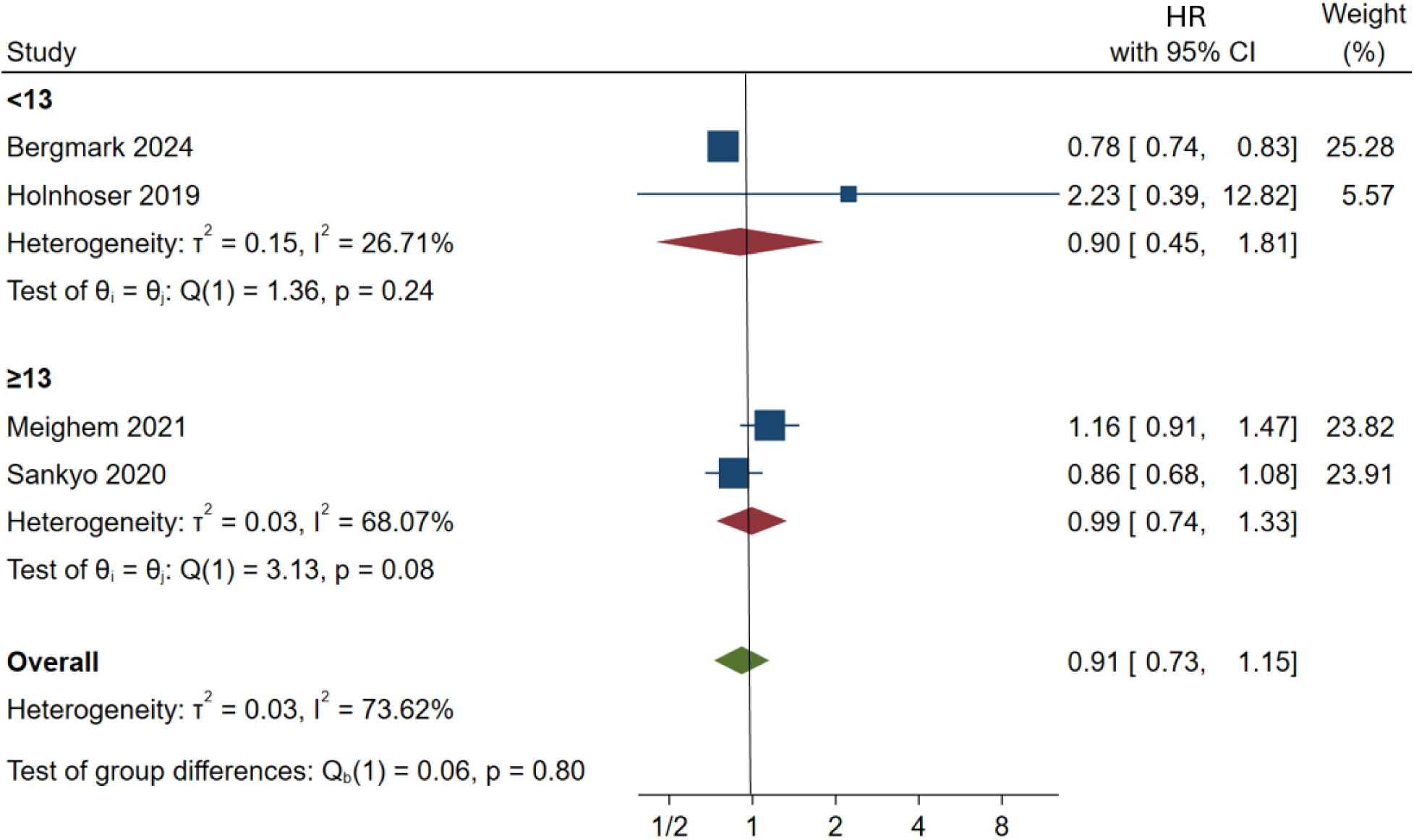
CRNM previous MI

### Risk of bias and Sensitivity analysis

As an evaluation of the risk of bias, Cochrane RoB tool was utilized (Supplementary Figure S1) and GRADE assessment was also conducted to enhance the clinicians in mapping the key decisions based on the evidence of outcomes as presented in Table 2. To overcome the problem of heterogeneity and the possibility of confounding variables, subgroup analyses were conducted according to the primary clinical variables: age, renal dysfunction, CHAUDSU-VASc score, heart failure, and history of myocardial infarction. The test as performed by Egger was also to study the strength of the results and presented in Supplementary Table S1 reveals that no small-study effects were obtained in studies.

**Table 2.**
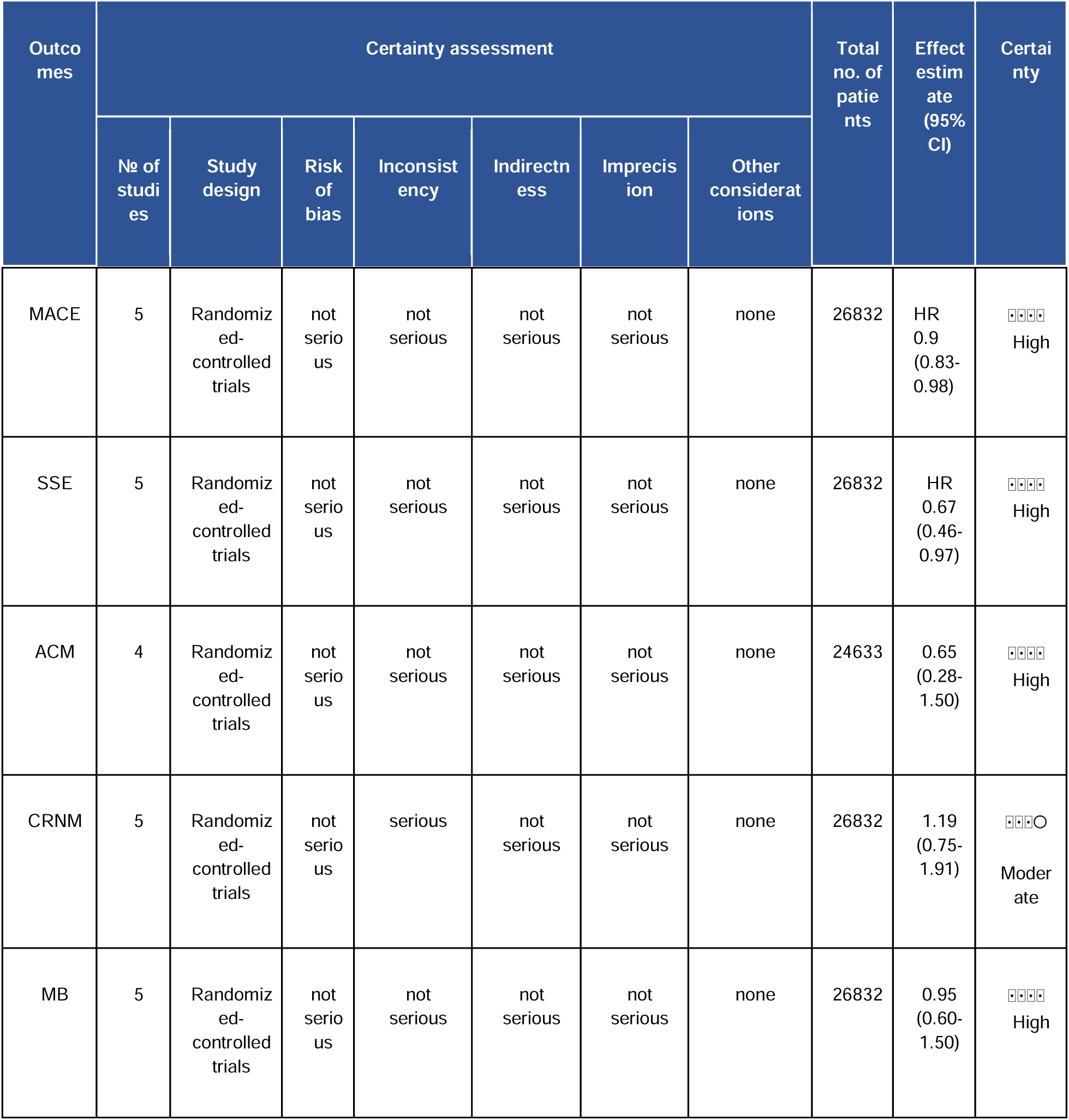
GRADE Assessment.

## Discussion

In this systematic review and meta-analysis, we provide a comprehensive evaluation of the efficacy and safety of edoxaban compared to warfarin in patients with NVAF as well as high-risk individuals who have undergone cardiac procedures such as PCI and TAVR. While edoxaban was associated with a reduced risk of major adverse cardiac events (MACE), stroke/systemic embolism (SSE), and major bleeding, warfarin was also a viable treatment choice in certain groups, especially those with a higher CHA 2D s VASc score, previous myocardial infarction (MI), and high heart failure load. These resultshighlight the importance of an individualized approach to anticoagulation treatment to balance thromboembolic protection and bleeding risks to maximize patient outcomes.

Patients with AF were at a much greater risk of MACE, SSE, and major bleeding, as they both have prothrombotic factors and cardiovascular factors. MACE, a term commonly used in observational studies and randomised controlled trials to evaluate the safety of cardiovascular events, comprises cardiovascular death, stroke, acute myocardial infarction (AMI), and revascularisation. [21] Our pooled analysis showed that edoxaban was superior to warfarin in the following areas. These results are consistent with the available literature. Xiangwen Liang et al. proved that edoxaban 30 and 60 mg have a significant impact on cardiovascular disease (CVD), major bleeding, and non-major bleeding events. Moreover, the incidences of stroke, MI, and adverse events (AEs) were not higher with higher doses (120 mg), which is further evidence of the need to optimize the dose in order to balance efficacy and safety. [11]

### Influence on Renal Function

Renal testing is necessary in the measurement of the anticoagulant effects since kidney failure can be a significant determinant of the drug excretion and therapy. Creatinine clearance (CrCl) is a rapid and inexpensive way of estimating renal function, with ranges of 90-120 mL/min considered normal in healthy young adults. [22]

In our analysis, renal function appeared to play a role in modulating clinical outcomes with edoxaban and warfarin, although findings varied across subgroups. In major bleeding, there was no notable difference between the two anticoagulants at different levels of CrCl. This differs from previous research, which indicated a safety benefit of edoxaban in patients with preserved renal function (CrCl ≥80 mL/min), owing to more efficient clearance of the drug. Although one study used slightly different CrCl categories (>95 and 50–90 mL/min), the overall conclusion that renal function affects pharmacokinetics is still valid. [23]

When considering the outcomes of efficacy, patients with preserved renal function were inclined to respond positively to edoxaban with reference to lowering major adverse cardiovascular events (MACE) and stroke/systemic embolism (SSE). Although the difference in MACE reduction by CrCl subgroup was not statistically significant, the overall trend favored better results with edoxaban in individuals with better renal function. This was better illustrated in the case of SSE, as the relative benefit of edoxaban appeared greater among patients with higher CrCl, indicating increased protective properties against thromboembolic occurrences in this group.

To conclude, these results indicate the possibility of renal function as a clinical variable in the process of anticoagulant choice. Moreover, in the literature, dose adjustment in cases of renal dysfunction has also been suggested; one study suggested edoxaban be lowered to 30 mg/day in case of moderate renal impairment (CrCl 30-50 mL/min) to prevent drug accumulation and maximum bleeding risk. [24] Although our meta-analysis considered different CrCl thresholds and failed to report statistically significant differences in the subgroups, the cumulative evidence was in support of the tailoring of anticoagulation according to the renal function of patients.

### High CHA□DS□-VASc Scores and Prior Heart Failure

In high-risk patients, specifically those with heart failure (HF) or with clinically relevant non-major bleeding (CRNM) because of having previous MI, or with a CHA□DS□-VASc vas score more than four, our study observed an association favoring warfarin over edoxaban. The CHA□DS□-VASc score is a guideline-based instrument that is used to evaluate the risk of thromboembolism and decide whether anticoagulation is required in patients with AF. [25] An increase in the score signifies the increased risk of thromboembolism, which requires anticoagulation treatment.

However, these findings are not fully supported by existing literature. As an example, Natalia S. Rost et al. found that in patients who had a history of an ischemic stroke or transient ischemic attack (IS/TIA), warfarin compared to high-dose edoxaban resulted in higher rates of intracranial hemorrhage per year. [26] Another analysis also indicated that edoxaban was more likely to prevent thromboembolic events among individuals with a higher CHA□DS□-VASc score [27] This difference between our findings and these studies may be attributable to differences in the study design, the population of patients, or the fact that the optimal dose has not been optimized in the high-risk populations. More observations should be placed under research to elucidate the same.

### Previous Exposure to Anticoagulants

A key finding in our study was a significant association favoring warfarin in patients previously exposed to vitamin K antagonists (VKA), and edoxaban in VKA-naive patients, defined as those with less than 60 days of continuous exposure to vitamin K antagonists before randomization. [28]

The results of the ENGAGE AF-TIMI 48 trial support this observation and show that low-dose edoxaban was not inferior to warfarin in VKA-naive patients (HR 0.92, 95% CI 0.73–1.15) but inferior to warfarin in VKA-experienced patients (HR 1.31, 95% CI 1.08–1.60; P interaction = 0.019). [29]

The efficacy of low-dose edoxaban and warfarin in stroke prevention is similar among VKA-naïve patients, presumably due to the absence of pre-existing coagulation adaptation. Chronic VKA treatment results in reduced levels of clotting factors II, VII, IX, and X, as warfarin inhibits vitamin K epoxide reductase (VKORC1) [30,31] Dependence on these anticoagulation mechanisms may therefore be increased in patients with long-term VKA exposure. Conversely, VKA-naïve patients lack such adaptations, and direct factor Xa inhibition by edoxaban can be similarly effective in thromboembolic prevention in this population.

### Influence of Age

Age subgroup analysis showed a significant difference in the efficacy and safety profile of edoxaban and warfarin. Edoxaban appeared to be more effective in younger patients (Less than 65 years) as the risk of stroke and systemic embolism (SSE) decreased significantly with the use of this intervention. Though the difference in Clinically Relevant Non-Major bleeding(CRNM) risk was not statistically significant between the two drugs, the trend still showed in favor of edoxaban.

Our results challenge earlier studies, such as Gentian Denas et al., which favored warfarin in younger patients.[32] This difference may be due to changing prescribing trends, better access to DOAC, or populations of patients. Younger people can also possess an improved renal clearance, fewer comorbidities, and improved adherence, which can be beneficial to the clinical efficacy of edoxaban.

On the contrary, anticoagulation is also influenced by the physiological changes in elderly population decisions. Frailty, characterized by reduced biological reserves and degraded homeostasis due to increasing age, is an essential element in deciding the anticoagulation in stroke prevention of patients who have atrial fibrillation (AF). [33] In frail elderly patients, such considerations have lead to warfarin being the preference as the its effects can be easily monitored and reversed compared to DOACs like edoxaban. [34,35]

### Strengths and limitations

This systematic review and meta-analysis encompasses an extensive comparison of the efficacy and safety of edoxaban and warfarin in different scenarios and groups of patients. Five clinical trials were included with a total of 26,832 participants reported in the trials highlights the generalizability and robustness of the findings. Addition of subgroup analyses aided in addressing heterogeneity and provide significant insight about particular groups of patients.

Using stringent and detailed tools for defining risk of bias (Cochrane RoB v2 tool and GRADE assessment) were utilized. Additionally, comparison of the effectiveness and safety of edoxaban and warfarin is achieved through the assessment of a number of essential outcomes (SSE, MACE, ACM, CRNM, and MB). This study highlights the practicality of individualized anticoagulation treatment, providing two-fold benefits, which includes assisting clinicians in making educated decisions to maximize patient outcomes and to reduce burden of preventable events.

The differences in the study designs, patient populations, and dose optimisations may be the reasons of dissonances and can affect the results comparison. Certain inferences, particularly those related to high-risk populations are not fully supported by the existent literature, and therefore, further studies are warranted in such cohorts. Although strong bias evaluation measures have been used, some inherent biases in the studies used persist, which may affect the overall results. Our study is based on the data of published studies, which might lead to publication bias and prevent the inclusion of unpublished but potentially relevant results.

## Conclusion

Edoxaban was associated with lower rates of major cardiac events, strokes, systemic embolisms, and major bleeding in non-valvular atrial fibrillation patients, especially those with a preserved kidney function and no prior warfarin exposure. However, warfarin maintains its ground for some underserved groups i.e. elderly patients. Personalizing anticoagulation therapy is essential to balance clot prevention with bleeding risks. Further studies are warranted to confirm our findings.

## Data Availability

All data produced in the present work are contained in the manuscript.

## DECLARATIONS

### Ethics approval and consent to participate

Not applicable

### Consent for publication

Not Applicable

### Availability of data and materials

The datasets supporting the conclusions of this article are included within the article and its additional files.

### Authors’ contributions

MAS, Conceptualization of draft, data extraction, formal analysis and manuscript writing. AM, data extraction, manuscript writing and reviewing. MWI, data screening, analysis and manuscript writing. MK, data screening and data extraction. SS, data analysis and manuscript writing. MHA, reviewing and editing the manuscript. AAK, revision of the manuscript.

## Acknowledgements

Not applicable

## SUPPLEMENTARY MATERIAL

**Figure S1.**
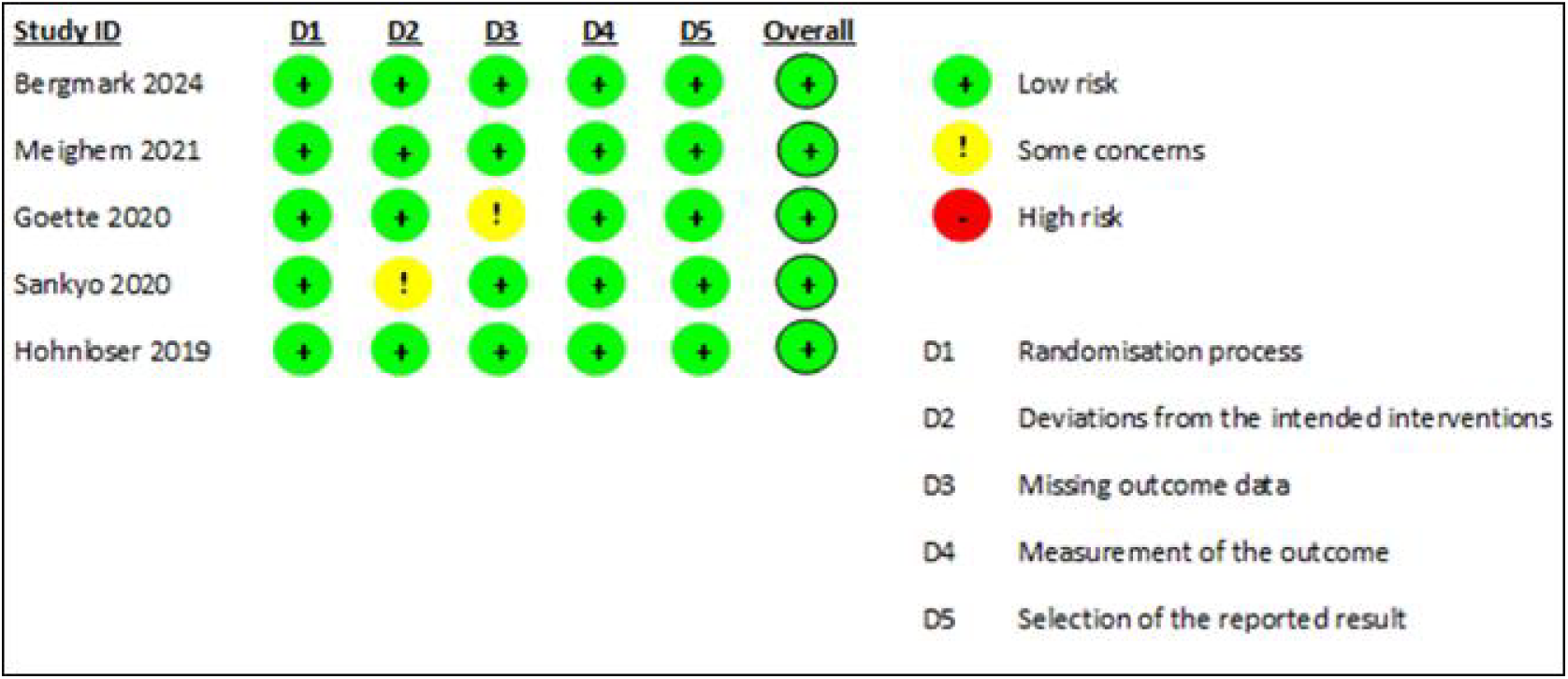
Cochrane Risk of bias assessment

---

**Table S1.**
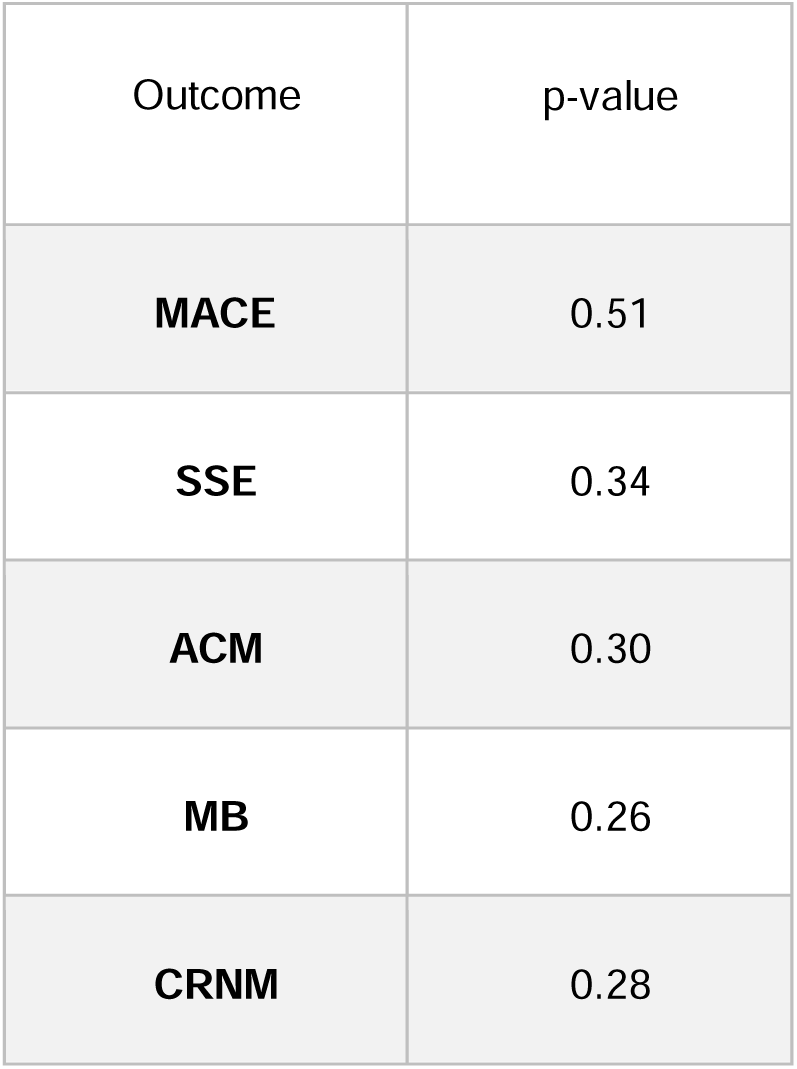
Egger’s test.

| Outcome | p-value |
| --- | --- |
| **MACE** | 0.51 |
| **SSE** | 0.34 |
| **ACM** | 0.30 |
| **MB** | 0.26 |
| **CRNM** | 0.28 |

## References

1. Ko D, Chung MK, Evans PT, Benjamin EJ, Helm RH. Atrial Fibrillation: A Review. JAMA. 2025 Jan 28;333(4):329–42.

2. Wu J, Zhang Y, Liao X, Lei Y. Anticoagulation Therapy for Non-valvular Atrial Fibrillation: A Mini-Review. Front Med. 2020 Jul 21;7.

3. Taduru S, Kamel H, Inohara T, Manning WJ, Piccini JP, Fonarow GC, et al. Trends in cardiovascular mortality related to atrial fibrillation in the United States, 2011 to 2018. J Am Heart Assoc. 2021;10(1):e020163. doi:10.1161/JAHA.120.020163

4. Zhou Z, Hu D, Chen J, et al. Understanding the causes of atrial fibrillation: a comprehensive review. BMC Public Health. 2022;22:14403. Available from: https://bmcpublichealth.biomedcentral.com/articles/10.1186/s12889-022-14403-2.

5. Fauchier L, Philippart R, Clementy N, Bourguignon T, Angoulvant D, Ivanes F, et al. How to define valvular atrial fibrillation? Archives of Cardiovascular Diseases. 2015 Oct;108(10):530–9.

6. Hamaguchi T, Iwanaga Y, Nakai M, Morita Y, Inoko M. Clinical Significance of Atrial Fibrillation Status in Patients With Percutaneous Coronary Intervention. CJC Open. 2021 Jul 6;3(11):1357–64.

7. Ryan T, Grindal A, Jinah R, Um KJ, Vadakken ME, Pandey A, et al. New-Onset Atrial Fibrillation After Transcatheter Aortic Valve Replacement. JACC: Cardiovascular Interventions. 2022 Mar;15(6):603–13.

8. Lee SY, Choi KH, Park TK, Kim J, Kim EK, Park SJ, et al. Impact of Atrial Fibrillation on Patients Undergoing Transcatheter Aortic Valve Implantation (TAVI): The K-TAVI Registry. Yonsei Med J. 2023 Jul;64(7):413–22.

9. Maan A, Khan MS, Khan SU, et al. A meta-analysis of the impact of pre-existing and new-onset atrial fibrillation on clinical outcomes in patients undergoing transcatheter aortic valve implantation. EuroIntervention. 2021;16(15):e1234–e1245.

10. Drikite L, Bedford JP, O’Bryan L, Petrinic T, Rajappan K, Doidge J, et al. Treatment strategies for new onset atrial fibrillation in patients treated on an intensive care unit: a systematic scoping review. Crit Care. 2021 Jul 21;25(1).

11. Kowalski M, Nowak J, Smith P, et al. Anticoagulation therapy in atrial fibrillation: The role of warfarin. Polish Heart Journal. 2022;79(3):123–130. Available from: https://journals.viamedica.pl/polish_heart_journal/article/view/98356.

12. Liu PH, Liu ZH, Niu MH, Chen P, Shi YB, He F, et al. A Comparative Study of the Clinical Benefits of Rivaroxaban and Warfarin in Patients With Non-valvular Atrial Fibrillation With High Bleeding Risk. Front Cardiovasc Med. 2022 Feb 16;9:803233.

13. Sonuga BO, Hellenberg DA, Cupido CS, Jaeger C. Profile and anticoagulation outcomes of patients on warfarin therapy in an urban hospital in Cape Town, South Africa. Afr J Prim Health Care Fam Med. 2016 May 31;8(1):e1–8.

14. Asinger RW, Shroff GR, Simegn MA, Herzog CA. Anticoagulation for Nonvalvular Atrial Fibrillation: Influence of Epidemiologic Trends and Clinical Practice Patterns on Risk Stratification and Net Clinical Benefit. Circ Cardiovasc Qual Outcomes. 2017 Sep;10(9):e003669.

15. Hohmann C, Lutz M, Vignali S, Borchert K, Seidel K, Braun S, et al. Clinical outcomes in patients receiving edoxaban or phenprocoumon for prevention of stroke in atrial fibrillation: a German real-world cohort study. Thrombosis J. 2022 Jul 4;20(1).

16. Zelniker TA, Ardissino M, Andreotti F, O’Donoghue ML, Yin O, Park JG, et al. Comparison of the Efficacy and Safety Outcomes of Edoxaban in 8040 Women Versus 13 065 Men With Atrial Fibrillation in the ENGAGE AF-TIMI 48 Trial. Circulation. 2021 Feb 16;143(7):673–84.

17. Lip GYH, Agnelli G. Edoxaban: a focused review of its clinical pharmacology. European Heart Journal. 2014 May 8;35(28):1844–55.

18. Sterne JAC, Savović J, Page MJ, Elbers RG, Blencowe NS, Boutron I, et al. RoB 2: a revised tool for assessing risk of bias in randomised trials. BMJ. 2019 Aug 28;l4898.

19. GRADEpro GDT: GRADEpro Guideline Development Tool [Software] McMaster University and Evidence Prime, 2025. Available from gradepro.org.

20. Higgins JPT. Measuring inconsistency in meta-analyses. BMJ. 2003 Sep 6;327(7414):557–60.

21. Bosco E, Hsueh L, McConeghy KW, Gravenstein S, Saade E. Major adverse cardiovascular event definitions used in observational analysis of administrative databases: a systematic review. BMC Med Res Methodol. 2021 Nov 6;21(1).

22. Shahbaz H, Rout P, Gupta M. Creatinine Clearance. In: StatPearls. Treasure Island (FL): StatPearls Publishing; 2024.

23. Wang Y, Li L, Wei Z, Lu S, Liu W, Zhang J, et al. Efficacy and Safety of Renal Function on Edoxaban Versus Warfarin for Atrial Fibrillation: A Systematic Review and Meta-Analysis. Medicines (Basel). 2023 Jan 16;10(1):13.

24. Kato T, Yamashita T, Sagara K, et al. Efficacy and safety of edoxaban compared with warfarin in patients with atrial fibrillation and heart failure in the ENGAGE AF-TIMI 48 trial. Eur J Heart Fail. 2016;18(9):1153–1161. Available from: 10.1002/ejhf.595.

25. Chen LY, Norby FL, Chamberlain AM, MacLehose RF, Bengtson LGS, Lutsey PL, et al. CHA_2_DS_2_VASc Score and Stroke Prediction in Atrial Fibrillation in Whites, Blacks, and Hispanics. Stroke. 2019 Jan;50(1):28–33.

26. Rost NS, Giugliano RP, Ruff CT, Murphy SA, Crompton AE, Norden AD, et al. Outcomes With Edoxaban Versus Warfarin in Patients With Previous Cerebrovascular Events: Findings From ENGAGE AF-TIMI 48 (Effective Anticoagulation With Factor Xa Next Generation in Atrial Fibrillation-Thrombolysis in Myocardial Infarction 48). Stroke. 2016 Aug;47(8):2075–82.

27. de Groot JR, Ruff CT, Murphy SA, Hamershock RA, Vehmeijer JT, Oude Ophuis AJM, et al. Edoxaban versus warfarin in patients with atrial fibrillation in relation to the risk of stroke: A secondary analysis of the ENGAGE AF-TIMI 48 study. Am Heart J. 2021 May;235:132–9.

28. Gencer B, Eisen A, Berger D, Nordio F, Murphy SA, Grip LT, et al. Edoxaban versus Warfarin in high-risk patients with atrial fibrillation: A comprehensive analysis of high-risk subgroups. American Heart Journal. 2022 May;247:24–32.

29. O’Donoghue ML, Ruff CT, Giugliano RP, Murphy SA, Grip LT, Mercuri MF, et al. Edoxaban vs. warfarin in vitamin K antagonist experienced and naive patients with atrial fibrillation†. European Heart Journal. 2015 Feb 16;36(23):1470–7.

30. Whirl-Carrillo M, Huddart R, Gong L, Sangkuhl K, Thorn CF, Whaley R, Klein TE. An Evidence-Based Framework for Evaluating Pharmacogenomics Knowledge for Personalized Medicine. Clin Pharmacol Ther. 2021;110(3):563–572. Available from: 10.1002/cpt.2350.

31. Hirsh J, Fuster V, Ansell J, Halperin JL. American Heart Association/American College of Cardiology Foundation Guide to Warfarin Therapy. Circulation. 2003 Apr;107(12):1692–711.

32. Denas G, Zoppellaro G, Granziera S, Pagliani L, Noventa F, Iliceto S, et al. Very Elderly Patients With Atrial Fibrillation Treated With Edoxaban: Impact of Frailty on Outcomes. JACC Adv. 2023 Aug 24;2(7):100569.

33. Bul M, Shaikh F, McDonagh J, Ferguson C. Frailty and oral anticoagulant prescription in adults with atrial fibrillation: A systematic review. Aging Med (Milton). 2022 Jun 1;6(2):195–206.

34. Zareh M, Davis A, Henderson S. Reversal of warfarin-induced hemorrhage in the emergency department. West J Emerg Med. 2011 Nov;12(4):386–92.

35. White K, Faruqi U, Cohen AAT. New agents for DOAC reversal: a practical management review. Br J Cardiol. 2022 Jan 12;29(1):1.

